# AutoMorph: Automated Retinal Vascular Morphology Quantification via a Deep Learning Pipeline

**DOI:** 10.1101/2022.05.26.22274795

**Authors:** Yukun Zhou, Siegfried K. Wagner, Mark Chia, An Zhao, Peter Woodward-Court, Moucheng Xu, Robbert Struyven, Daniel C. Alexander, Pearse A. Keane

## Abstract

**Purpose:** To externally validate a deep learning pipeline (AutoMorph) for automated analysis of retinal vascular morphology on fundus photographs. AutoMorph has been made publicly available (https://github.com/rmaphoh/AutoMorph), facilitating widespread research in ophthalmic and systemic diseases.

**Methods:** AutoMorph consists of four functional modules: image pre-processing, image quality grading, anatomical segmentation (including binary vessel, artery/vein, and optic disc/cup segmentation), and vascular morphology feature measurement. Image quality grading and anatomical segmentation use the most recent deep learning techniques. We employ a model ensemble strategy to achieve robust results and analyse the prediction confidence to rectify false gradable cases in image quality grading. We externally validate each module’s performance on several independent publicly available datasets.

**Results:** The EfficientNet-b4 architecture used in the image grading module achieves comparable performance to the state-of-the-art for EyePACS-Q, with an F1-score of 0.86. The confidence analysis reduces the number of images incorrectly assessed as gradable by 76%. Binary vessel segmentation achieves an F1-score of 0.73 on AV-WIDE and 0.78 on DR-HAGIS. Artery/vein scores 0.66 on IOSTAR-AV, and disc segmentation achieves 0.94 in IDRID. Vascular morphology features measured from AutoMorph segmentation map and expert annotation show good to excellent agreement.

**Conclusions:** AutoMorph modules perform well even when external validation data shows domain differences from training data, e.g., with different imaging devices. This fully automated pipeline can thus allow detailed, efficient and comprehensive analysis of retinal vascular morphology on colour fundus photographs.

**Translational Relevance:** By making AutoMorph publicly available and open source, we hope to facilitate ophthalmic and systemic disease research, particularly in the emerging field of ‘oculomics’.

## Introduction

The widespread availability of rapid, non-invasive retinal imaging has been one of the most notable developments within ophthalmology in recent decades. The significance of the retinal vasculature for assessing ophthalmic disease is well known, however there is also growing interest in its capacity to provide valuable insights into systemic disease, a field which has been termed ‘oculomics.’^1–4^. Narrowing of the retinal arteries is associated with hypertension and atherosclerosis ^5–8^, whilst dilation of the retinal veins is linked with diabetic retinopathy ^9–11^. Increased tortuosity of the retinal arteries is also associated with hypercholesterolaemia and hypertension ^12–14^. Considering that manual vessel segmentation and feature extraction can be extremely time consuming, as well as poorly reproducible ^15^, there has been growing interest in the development of tools which can extract retinal vascular features in a fully-automated manner.

In recent decades, a large body of technical work has focused on retinal vessel map segmentation. Performance has improved dramatically through employing a range of techniques from unsupervised graph- and feature-based methods ^16–20^ to supervised deep learning models ^21^. Despite this progress, the widespread use of these techniques in clinical research has been limited by a number of factors. Firstly, technical papers ^21–25^ often focus on performing a single function whilst ignoring upstream and downstream tasks, such as pre-processing ^24, 25^ and feature measurement ^21–23^. Secondly, existing techniques often perform poorly when applied to real-world clinical settings limited by poor generalisability outside of the environment in which they were developed ^26, 27^.

Although some software has been utilised for clinical research, most are only semi-automated, requiring human intervention for correcting vessel segmentation and artery/vein identification ^6, 24, 25, 28, 29^. This limits process efficiency and introduces subjective bias, potentially influencing the final outcomes. Further, most existing software has not integrated the crucial functions needed for such a pipeline, namely image cropping, quality assessment, segmentation, and vascular feature measurement. For example, poor quality images in research cohorts often need to be manually filtered by physicians, which generates a considerable workload. There is also the potential to improve the performance of underlying segmentation algorithms by employing the most recent advances in machine learning, thus enhancing the accuracy of vascular feature measurements.

In this study, we explore the feasibility of a deep learning pipeline providing automated analysis of retinal vascular morphology from colour fundus photographs. We highlight three unique advantages of the proposed “AutoMorph” pipeline:

1. AutoMorph consists of four functional modules, including: (a) retinal image pre-processing, (b) image quality grading, (c) anatomical segmentation (binary vessel segmentation, artery/vein segmentation, and optic disc segmentation), and (d) morphological feature measurement.
2. AutoMorph alleviates the need for physician intervention by addressing two key areas. Firstly, we employ an ensemble technique with confidence analysis to reduce the number of ungradable images which are incorrectly classified as being gradable (false gradable images). Secondly, accurate binary vessel segmentation and artery/vein identification reduce the need for manual rectification.
3. AutoMorph generates a diverse catalogue of retinal feature measurements that previous work indicates has the potential to be used for the exploration of ocular biomarkers for systemic disease.

Perhaps most importantly, we made AutoMorph publicly available with a view to stimulating breakthroughs in the emerging field of ‘oculomics’.

## Methods

The AutoMorph pipeline consists of four modules: 1) image pre-processing, 2) image quality grading, 3) anatomical segmentation, and 4) metric measurement (Figure 1). Source code for this pipeline is available from (https://github.com/rmaphoh/AutoMorph).

**Figure 1.**
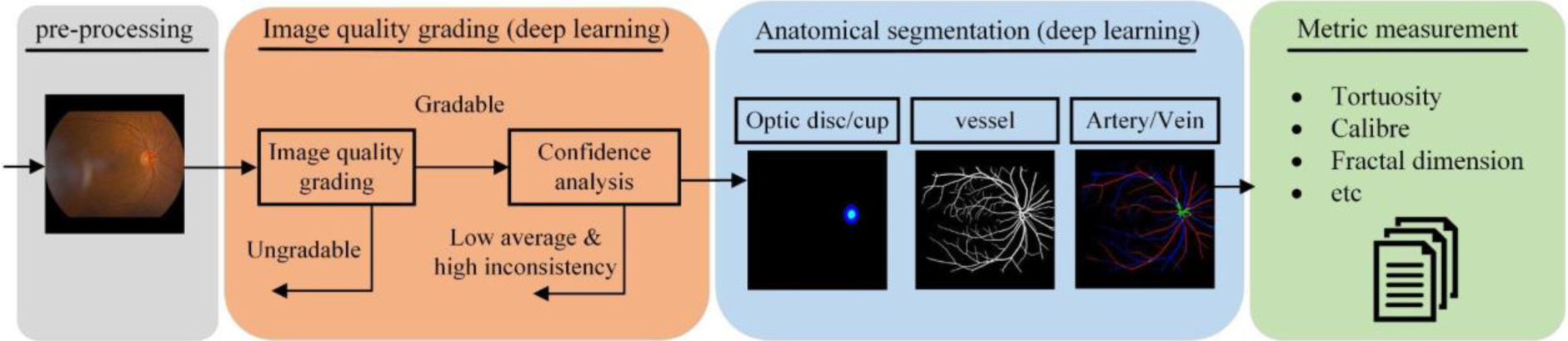
Diagram of the proposed AutoMorph pipeline. The input is colour fundus photography and the final output is the measured vascular morphology features. Image quality grading and anatomical segmentation modules use deep learning models. Confidence analysis decreases false gradable images in the image quality grading module.

## Datasets

The datasets used for development and external validation of the deep learning models described in this work are summarised in Table 1 and Supplementary Material S1. For model training, we chose publicly available datasets which contain a large quantity of annotated images ^30^. Importantly, a diverse combination of public datasets was used in order to enhance external generalisability. To validate the models, we externally evaluated the performance of those trained models on datasets distinct from those on which they were trained (e.g., different imaging devices, country of origin, and types of pathology). All the datasets provide the retinal fundus photographs and the corresponding expert annotation. For image quality grading datasets, using EyePACS-Q as an example, two experts grade each image into three categories: good, usable, and reject quality, determined by image illumination, artefacts, and the diagnosability of the general eye diseases to the experts. For anatomical segmentation datasets, such as the DRIVE dataset for binary vessel segmentation task, two experts annotate each pixel as vessel or background, thus generating a ground truth map with the same size of the retinal fundus photographs, where white colour indicates vessel pixels and black colour for the background. More details can be referred to Supplementary Material S1.

**Table 1.**
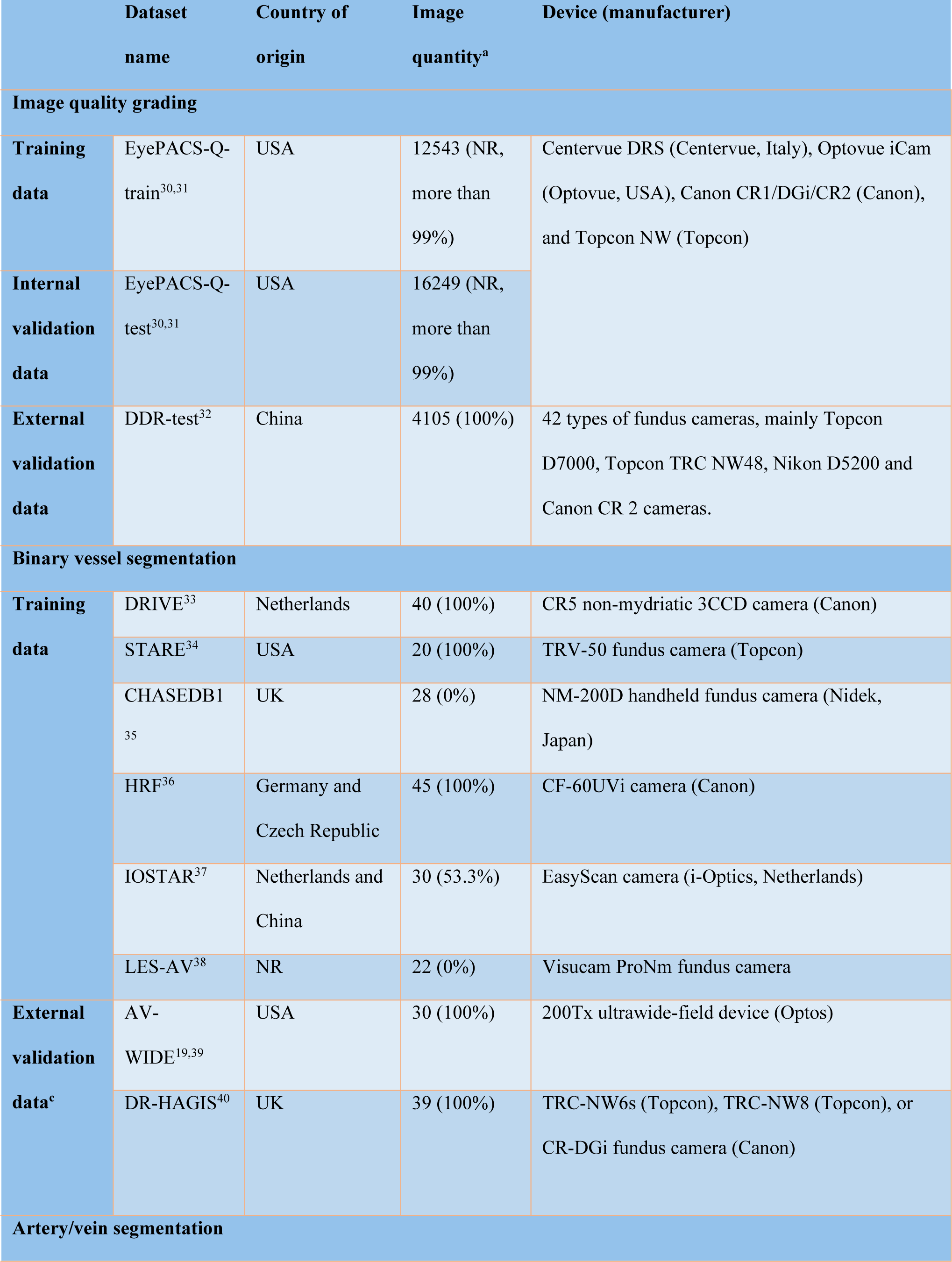
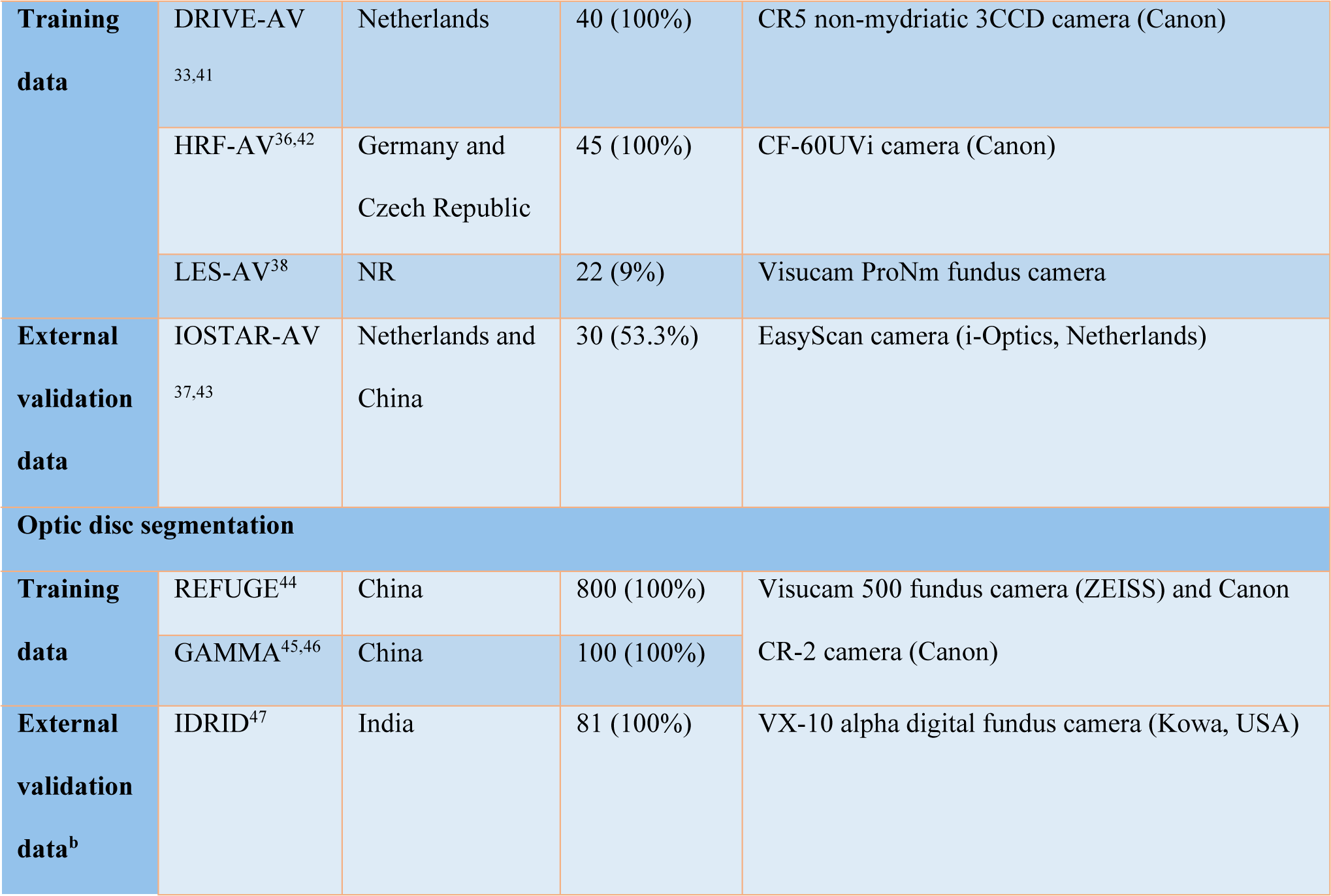
Characteristics of the training and external validation data. External validation data is unseen for model training and purely used to evaluate the trained model performance on out-of-distribution data with different countries of origin and imaging devices. EyePACS-Q, a subset of EyePACS with image quality grading annotation. a, image quantity indicates the image number used in this work and the brackets list the proportion of macula-centred images. b, evaluating on disc due to no cup annotation. c, although we have evaluated the binary vessel segmentation model on ultra-widefield retinal fundus dataset AV-WIDE, we recommend using AutoMorph on retinal fundus photographs with a 25-60 field of view (FOV), as all the deep learning models are trained using images with FOV equals to 25-60 degree, and the pre-processing step is tailored for images with this FOV. NR, not reported.

## Modules

### Image pre-processing

Retinal fundus photographs often contain superfluous background, resulting in dimensions that deviate from a geometric square. To account for this, we employed a technique that combines thresholding, morphological image operations, and cropping ^31^ to remove the background so that the resulting image conforms to a geometric square (examples shown in Supplementary Material S2).

### Image quality grading

To filter out ungradable images which often fail in segmentation and measurement modules, AutoMorph incorporates a classification model to identify ungradable images. The model classifies each image as good, usable, or reject quality. In our study, good and usable images were considered to be gradable, however this decision may be modified in scenarios where there is sufficient data to include only good quality images. We employed EfficientNet-B4 ^48^ as the model architecture and performed transfer learning on EyePACS-Q. Further details are outlined in Supplementary Material S2.

### Anatomical segmentation

Vascular structure is thin and elusive especially against low-contrast backgrounds. To enhance binary vessel segmentation performance, AutoMorph uses an adversarial segmentation network ^23^. Six public datasets were used for model training (Table 1). Accurate artery/vein segmentation is a long-standing challenge. To address this, we employed an information fusion network ^22^ which is tailored for artery/vein segmentation. Three datasets were used for training. Parapapillary atrophic changes, which can be a hallmark of myopia or glaucoma, can cause large errors in disc localisation and segmentation. To counter this, AutoMorph employs a coarse-to-fine deep learning network ^49^, which has achieved first place for disc segmentation in the MICCAI 2021 GAMMA challenge ^45, 46^. Two public datasets were utilised in model training. Further detailed information is provided in Supplementary Material S3.

### Vascular morphology feature measurement

AutoMorph measures a series of clinically-relevant vascular features, as summarised in Figure 2 (comprehensive list in Supplementary Material S5). Three different calculation methods for vessel tortuosity are provided including distance measurement tortuosity, squared curvature tortuosity ^50^, and tortuosity density ^51^. The fractal dimension value (Minkowski-Bouligand dimension) ^52^ provides a measurement of vessel complexity. The vessel density indicates the ratio between the area of vessels to the whole image. For vessel calibre, AutoMorph calculates the central retinal arteriolar equivalent (CRAE) and central retinal venular equivalent (CRVE), as well as the arteriolar-venular ratio (AVR) ^53–55^. AutoMorph measures the features in standard regions, including ZONE B (the annulus 0.5-1 optic disc diameter from the disc margin) and ZONE C (the annulus 0.5-2 optic disc diameter from the disc margin) ^29^. Considering ZONE B and ZONE C of macular-centred images may be out of the circular fundus, the features for the whole image are also measured.

**Figure 2.**
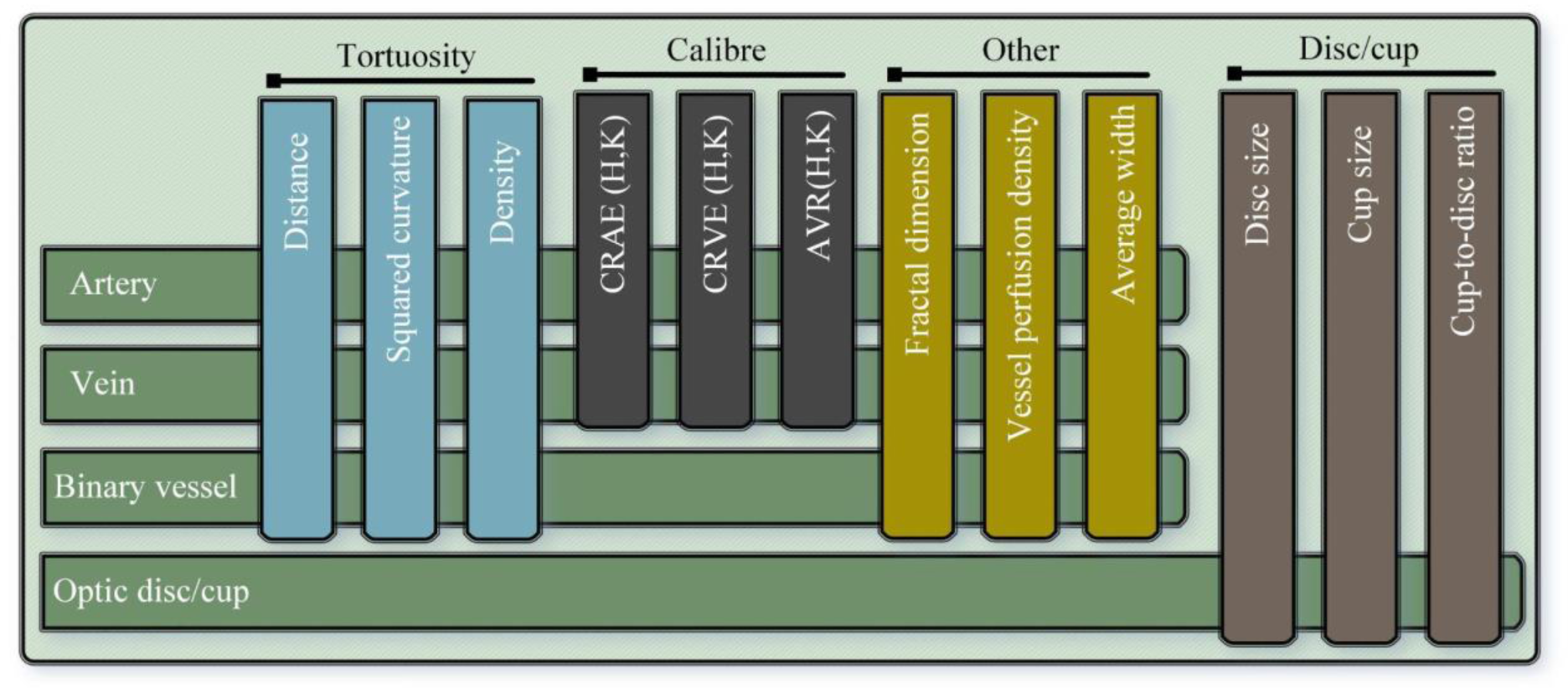
Features measured by AutoMorph, including tortuosity, vessel calibre, disc/cup, and others. For each image, the optic disc/cup information is measured, including the height and width, as well as cup-to-disc ratio. For binary vessels, the tortuosity, fractal dimension, vessel density, and average width are measured. Besides these features, arteries/veins are also used for measuring calibre features CRAE, CRVE, and AVR by Hubbard and Knudtson methods.

### Ensemble and confidence analysis

In model training, 80% of training data is for model training and 20% of it is used to tune the training hyperparameters, such as scheduling the learning rate. In retinal image grading, we ensemble the output from eight trained models with different subsets of training data, as it generally gives a more robust result ^56^. Moreover, the average value and standard deviation (SD) of the eight possibilities are calculated for confidence analysis. Average probability indicates the average confidence of prediction. Low average cases are prone to false predictions, such as Figure 3(c). Meanwhile, SD represents the inconsistency between models. High inconsistency likely corresponds to a false prediction, as shown in Figure 3(d). The images with either low average probability or high SD are automatically recognised as low confidence images and rectified as ungradable. False gradable images can fail the anatomical segmentation module thus generating a large error in vascular feature measurement. The confidence analysis economises physician intervention and increases the reliability of AutoMorph. To our knowledge, this is the first report of a confidence analysis combined with the model ensemble integrated within the vessel analysis pipeline. An average threshold corresponds to a change of operating point and SD threshold involved in uncertainty theory. In this work, we set an average threshold of 0.75 and a SD threshold of 0.1 to filter out false gradable images. Specifically, the average probability lower than 0.75 or SD larger than 0.1 will be rectified as ungradable images. The rationale for selecting threshold values is based on the probability distribution histogram on tuning data. More details are described in Supplementary Material S2.

**Figure 3.**
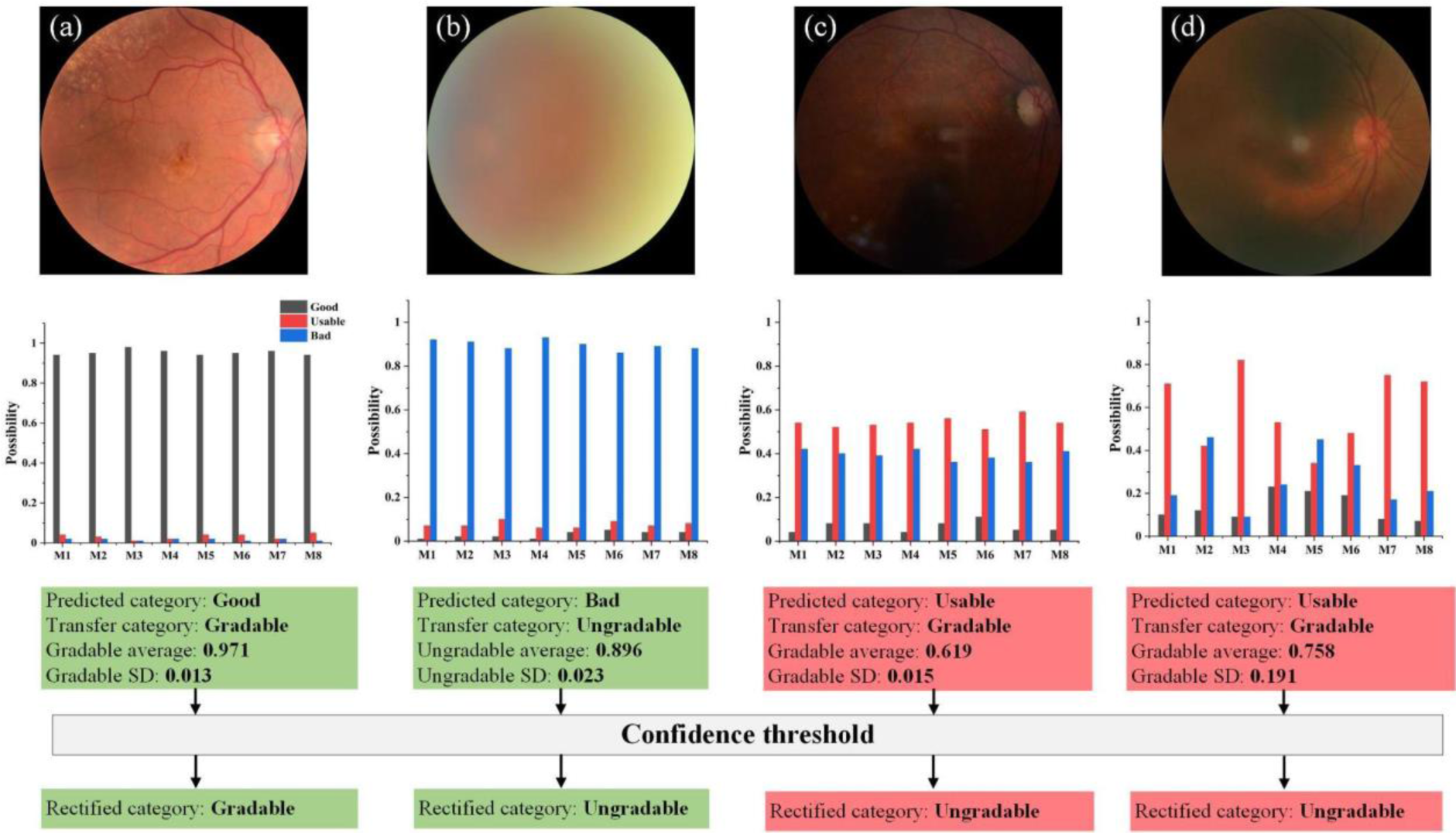
Confidence analysis for image quality grading. M1-M8 represents the eight ensemble models. For each image, the predicted category is transferred as gradable or ungradable (good and usable are as gradable, reject as ungradable). The average probability and SD are calculated for the predicted category. (a) and (b) are two image cases with high confidence in prediction. The case (c) is classified as gradable quality in low average probability of 0.619 while (d) is in high SD 0.191, which are defined as low confidence images in our work. Although (c) and (d) are preliminarily classified as gradable, the final classification is rectified as ungradable with the confidence threshold.

### Statistical analyses and compared methods

For deep learning functional modules, the well-established expert annotation is used as reference standard to quantitatively evaluate the module performance. We calculated Sensitivity, Specificity, Positive predictive value (Precision), Accuracy, Area-Under-Curve Receiver Operating Characteristic (AUC-ROC), F1-score, and IoU metrics to verify the model performance. These metric definitions are

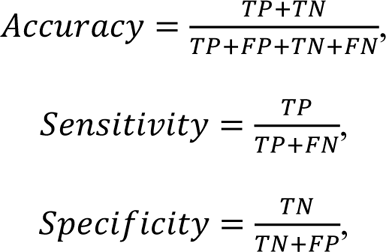

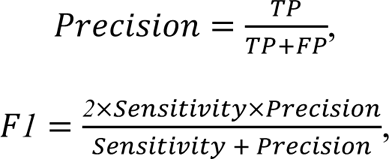

where TP, TN, FP, and FN respectively indicate true positive, true negative, false positive, and false negative. AUC-ROC curve is a performance measurement for classification problems at various threshold settings. It tells how much the model is capable of distinguishing between classes. In segmentation tasks, Intersection of Union (IoU) measures the overlap degree between ground truth maps and segmentation maps. Following the same setting ^31, 39, 57–59^, we set the ungradable images as the positive class in image quality grading. The probability of the ungradable category equals that of reject quality, and the probability of the gradable category is the sum of good quality and usable quality. As introduced in confidence analysis, we use a mean value of 0.75 and SD of 0.1 as thresholds to get the final rectified gradable and ungradable categories. For binary vessel segmentation, each pixel of the retinal fundus photograph corresponds to a binary classification task. The vessel pixel is positive class and the background pixel is negative. The probability range for each pixel is from 0 to 1, where a larger value indicates a higher probability of being a vessel pixel. We threshold the segmentation map with 0.5 which is a standard threshold for binary medical image segmentation tasks. Optic disc segmentation is similar to binary vessel segmentation, the difference is that the positive class is the optic disc pixel. For artery/vein segmentation, each pixel has a four-class probability of artery, vein, uncertain pixel, and background. Following standard settings for multi-class segmentation tasks, the category with the largest probability across the four classes is the thresholded pixel category. More information is listed in Supplementary Material S3.

We conducted the quantitative comparison to other competitive methods to characterise the generalisability of AutoMorph using external validation. We used internal validation results from other published work to provide a benchmark for a well-performing model. These methods used a reasonable proportion of data for model training and the remainder for internal validation (e.g., five-fold validation that means 80% of images are used for training and tuning, while 20% are used for validating the trained model), and claimed that they have achieved state-of-the-art performance. As introduced in Table 1, the models of AutoMorph are trained on several public datasets and externally validated on separate datasets, while the compared methods ^39, 57–59^ are trained in the same domain data as the validation data, but with fewer training images. The goal of the comparison was not to prove the technical strengths of AutoMorph over recent methods, since this has already been verified in previously published work ^22, 23, 47, 48^. Rather, we aim to demonstrate that due to the diversity of its training data, Automorph performs well on external datasets, even when these datasets include pathology and show large domain differences from the training data. Additionally, to compare method’s technical superiority, we have provided the internal validation of AutoMorph in Supplementary Material S3.

Considering we employ standard formulas ^29, 50–52^ to measure vascular morphology features, the measurement error only comes from inaccuracy of anatomical segmentation. In order to evaluate measurement error that occurs as a result of vessel segmentation, we respectively measure the vascular features based on AutoMorph segmentation and expert vessel annotation, and then draw Bland-Altman plots. Following the same evaluation ^3, 60^, intraclass correlation coefficients (ICC) are calculated to quantitatively show agreement. Additionally, the boxplots of difference between the vascular features from AutoMorph segmentation and expert annotation are listed in Supplementary Material S4.

## Results

Results for external validation of AutoMorph are summarized in Table 2.

**Table 2.**
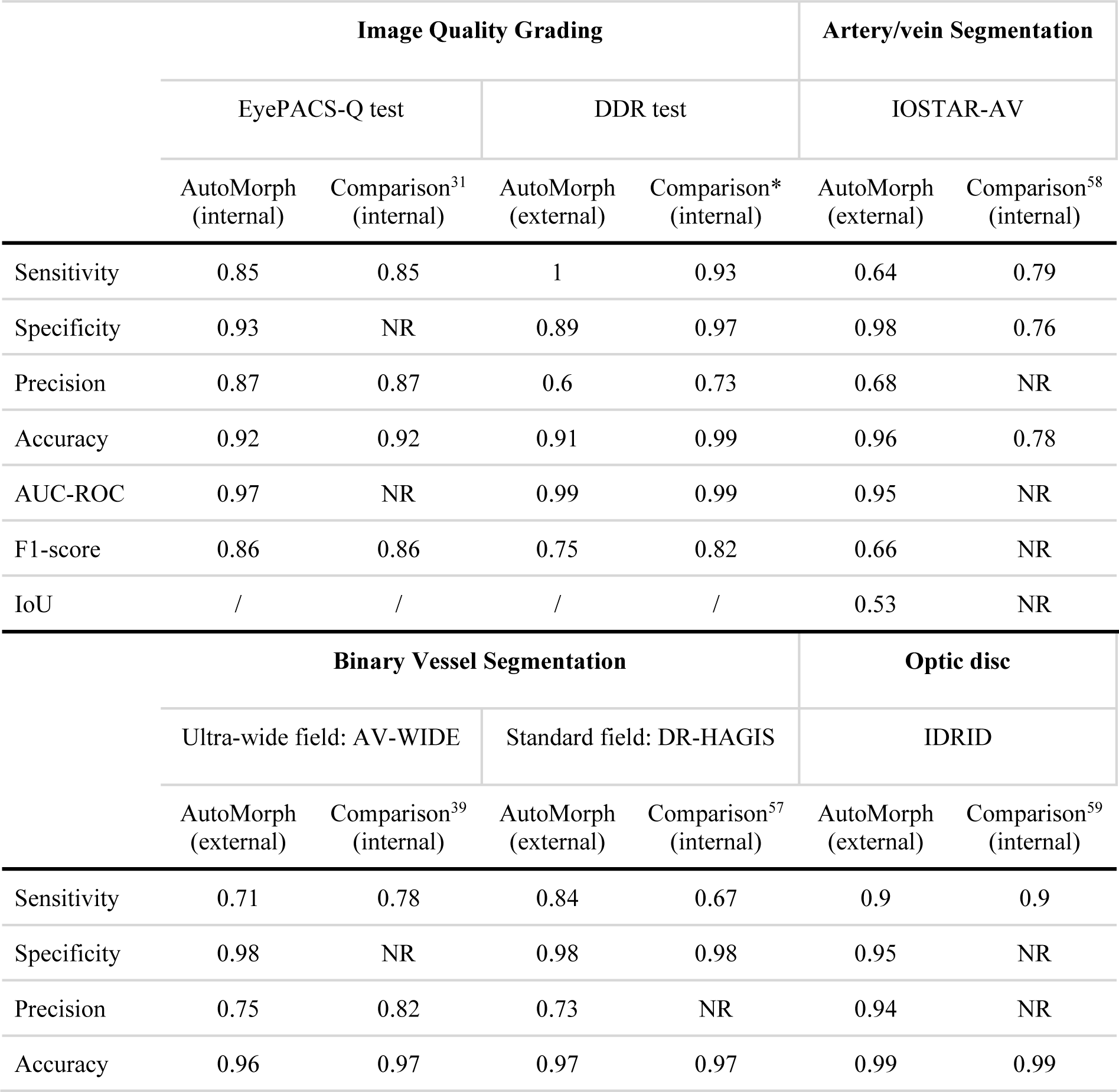
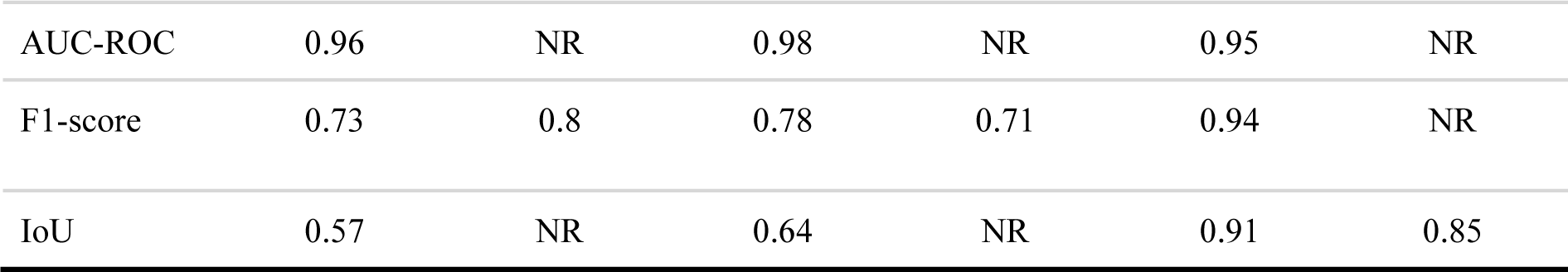
Validation of functional modules and comparison with other methods. NR, not reported. “Internal” indicates the validation and training data are from the same dataset but isolated. “External’’ means that validation data is from external datasets. The comparisons are with competitive methods of image quality grading ^31^, binary vessel segmentation ^39, 57^, artery/vein segmentation ^58^, and optic disc segmentation ^59^. * Due to no comparison method on the DDR test, we compare AutoMorph (external) to the same architecture EfficientNet-b4 which is trained with DDR train data (internal).

|  | Image Quality Grading |  |  |  | Artery/vein Segmentation |  |
| --- | --- | --- | --- | --- | --- | --- |
|  | EyePACS-Q test |  | DDR test |  | IOSTAR-AV |  |
|  | AutoMorph (internal) | Comparison<sup>31</sup> (internal) | AutoMorph (external) | Comparison* (internal) | AutoMorph (external) | Comparison<sup>58</sup> (internal) |
| Sensitivity | 0.85 | 0.85 | 1 | 0.93 | 0.64 | 0.79 |
| Specificity | 0.93 | NR | 0.89 | 0.97 | 0.98 | 0.76 |
| Precision | 0.87 | 0.87 | 0.6 | 0.73 | 0.68 | NR |
| Accuracy | 0.92 | 0.92 | 0.91 | 0.99 | 0.96 | 0.78 |
| AUC-ROC | 0.97 | NR | 0.99 | 0.99 | 0.95 | NR |
| F1-score | 0.86 | 0.86 | 0.75 | 0.82 | 0.66 | NR |
| IoU | / | / | / | / | 0.53 | NR |
|  | Binary Vessel Segmentation |  |  |  | Optic disc |  |
|  | Ultra-wide field: AV-WIDE |  | Standard field: DR-HAGIS |  | IDRID |  |
|  | AutoMorph (external) | Comparison<sup>39</sup> (internal) | AutoMorph (external) | Comparison<sup>57</sup> (internal) | AutoMorph (external) | Comparison<sup>59</sup> (internal) |
| Sensitivity | 0.71 | 0.78 | 0.84 | 0.67 | 0.9 | 0.9 |
| Specificity | 0.98 | NR | 0.98 | 0.98 | 0.95 | NR |
| Precision | 0.75 | 0.82 | 0.73 | NR | 0.94 | NR |
| Accuracy | 0.96 | 0.97 | 0.97 | 0.97 | 0.99 | 0.99 |
| AUC-ROC | 0.96 | NR | 0.98 | NR | 0.95 | NR |
| F1-score | 0.73 | 0.8 | 0.78 | 0.71 | 0.94 | NR |
| IoU | 0.57 | NR | 0.64 | NR | 0.91 | 0.85 |

### Image quality grading

The internal validation is on EyePACS-Q test data. For fair comparison to method ^31^, we evaluate the image quality grading performance of categorising good, usable, and reject quality. The quantitative results are listed in Table 2. The classification F1-score has achieved 0.86, in par with the state-of-the-art method with F1-score of 0.86 ^31^. After transferring the prediction to gradable (good and usable quality) and ungradable (reject quality), the confusion matrix of validation on the EyePACS-Q test is listed in Figure 4. We can learn that the confidence thresholding brings a trade-off in performance metrics, suppressing false gradable ratio but simultaneously increasing false negative. False gradable images are prone to fail the anatomical segmentation module and generate large errors and outliers in vascular feature measurement. Although this thresholding filters out some adequate quality images, it maintains the reliability of AutoMorph.

**Figure 4.**
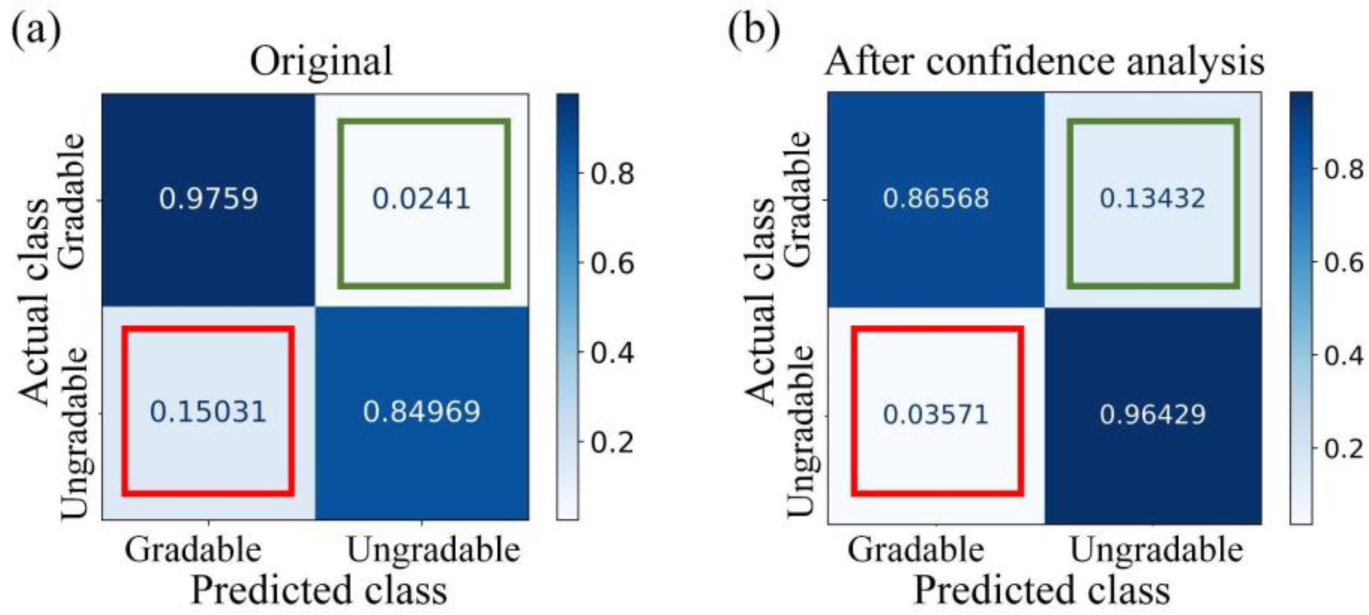
The confusion matrix of the grading results on EyePACS-Q test data. (a) shows the results before confidence thresholding while (b) is the results after it. The value is normalised in rows. The diagonal includes the correct classification ratio, while the red box indicates false gradable (i.e., ungradable images are wrongly classified as gradable) and the green box shows the percentage of false ungradable (i.e., gradable images are wrongly categorised as ungradable). The false gradable of (b) is reduced by 76.2% compared with that of (a), while the false ungradable increases in (b).

The external validation is on the DDR test data. As DDR only includes two categories in image quality annotation – gradable and ungradable, we firstly transfer the AutoMorph prediction of good and usable quality as gradable, and reject quality as ungradable, and then evaluate the quantitative results. Although the difference in the annotation might underestimate the AutoMorph image quality grading capability, the performance is satisfactory compared to the internal group, as shown in Table 2. The confusion matrix and AUC-ROC curve are shown in Supplementary Material S2. All ungradable images are correctly identified, which is significant for AutoMorph’s reliability.

### Anatomical segmentation

Visualisation results are presented in Figure 5 and quantitative results are listed in Table 2. For binary vessel segmentation, the two public datasets AV-WIDE and DR-HAGIS are employed in model validation. The binary vessel segmentation model works comparably to SOTA performance on the fundus photography data (DR-HAGIS) and moderately so on ultra-widefield data (AV-WIDE). For artery/vein segmentation, the performance is validated on the IOSTAR-AV dataset. Compared with the most recent method ^58^, AutoMorph achieves lower sensitivity but much higher specificity. The visualisation results of two challenging cases from Moorfields Eye Hospital and ORIGA are shown in Supplementary Material S3. For optic disc segmentation, we validated the performance on the dataset IDRID. The performance is on the par with the compared method ^59^ and the F1-score is slightly higher. Although pathology disturbs, the segmentation disc shows robustness.

**Figure 5.**
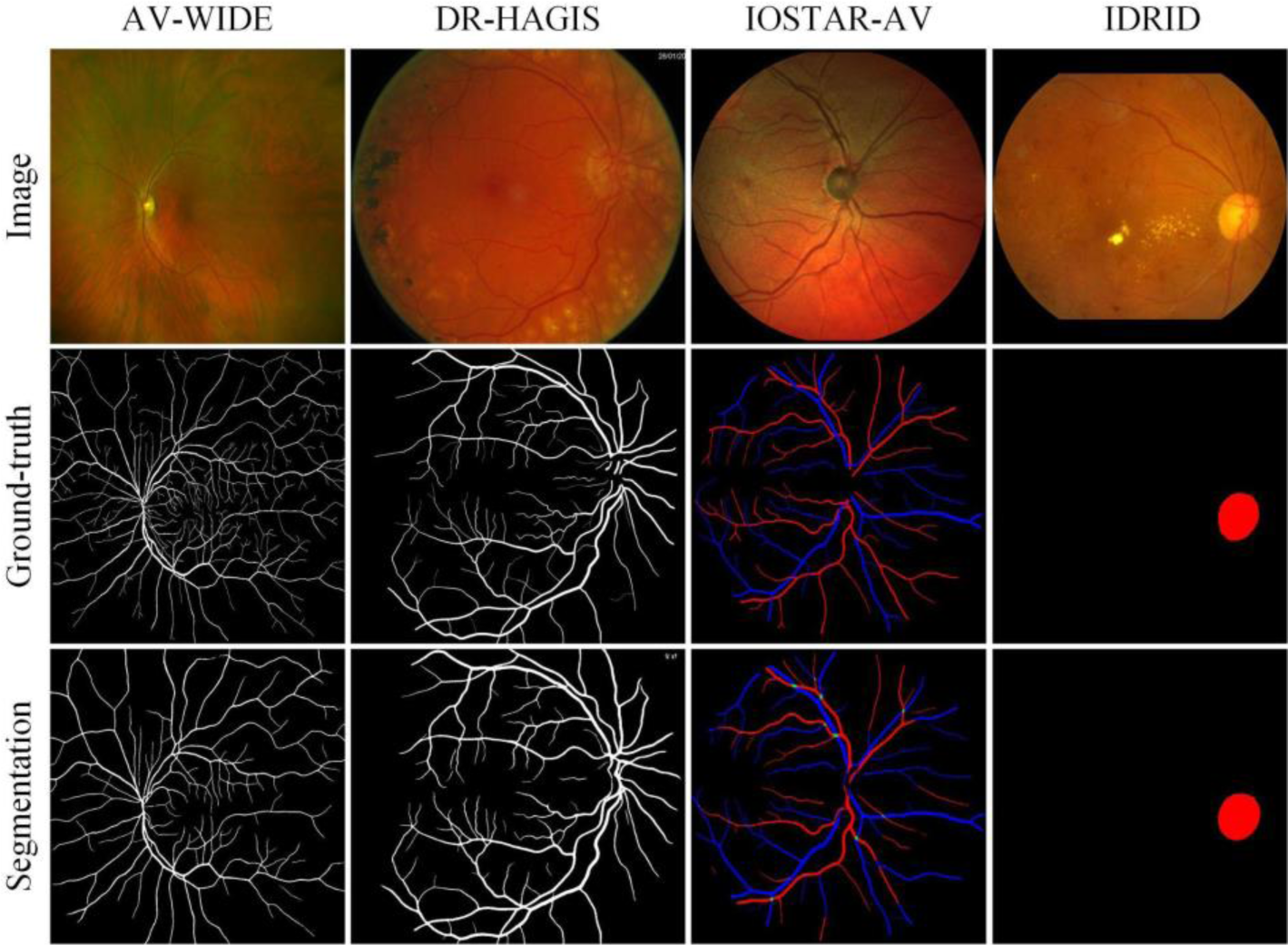
Visualisation results of anatomical segmentation, including binary vessel (first two columns), artery/vein (third column), and optic disc (final column).

### Vascular feature measurement

The ICCs between AutoMorph features and expert features are listed in Table 3. For binary vessel morphology, the fractal dimension, vessel density, and average width metrics all achieve excellent reliability (ICC>0.9). The other metrics show good consistency. Bland-Altman plots for ZONE B are shown in Figure 6. All features show agreement (Fractal dimension: mean difference (MD) is -0.01 and 95% limits of agreement (LOA) is -0.05 to 0.03; Vessel density: MD 0.001 and LOA from 0 to 0.002; Average width: MD 1.32 pixels and LOA from 0.44 to 2.19; Distance tortuosity: MD 0.02 and LOA from -2.18 to 2.22; Squared curvature tortuosity: MD -1.02 and LOA from -14.59 to 12.56; Tortuosity density: MD 0.02 and LOA from -0.09 to 0.13; CRAE Hubbard: MD -0.13 and LOA from -2.49 to 2.24; CRVE Hubbard: MD 0 and LOA from -2.9 to 2.9; AVR Hubbard: MD -0.03 and LOA from -0.17 to 0.11). The results at ZONE C and the whole image are listed in Supplementary Material S4. Note that for the metrics CRAE, CRVE, and average width, measurements are presented in pixels, as resolution information is unknown. Some images with large errors are listed in Supplementary Material S4.

**Figure 6.**
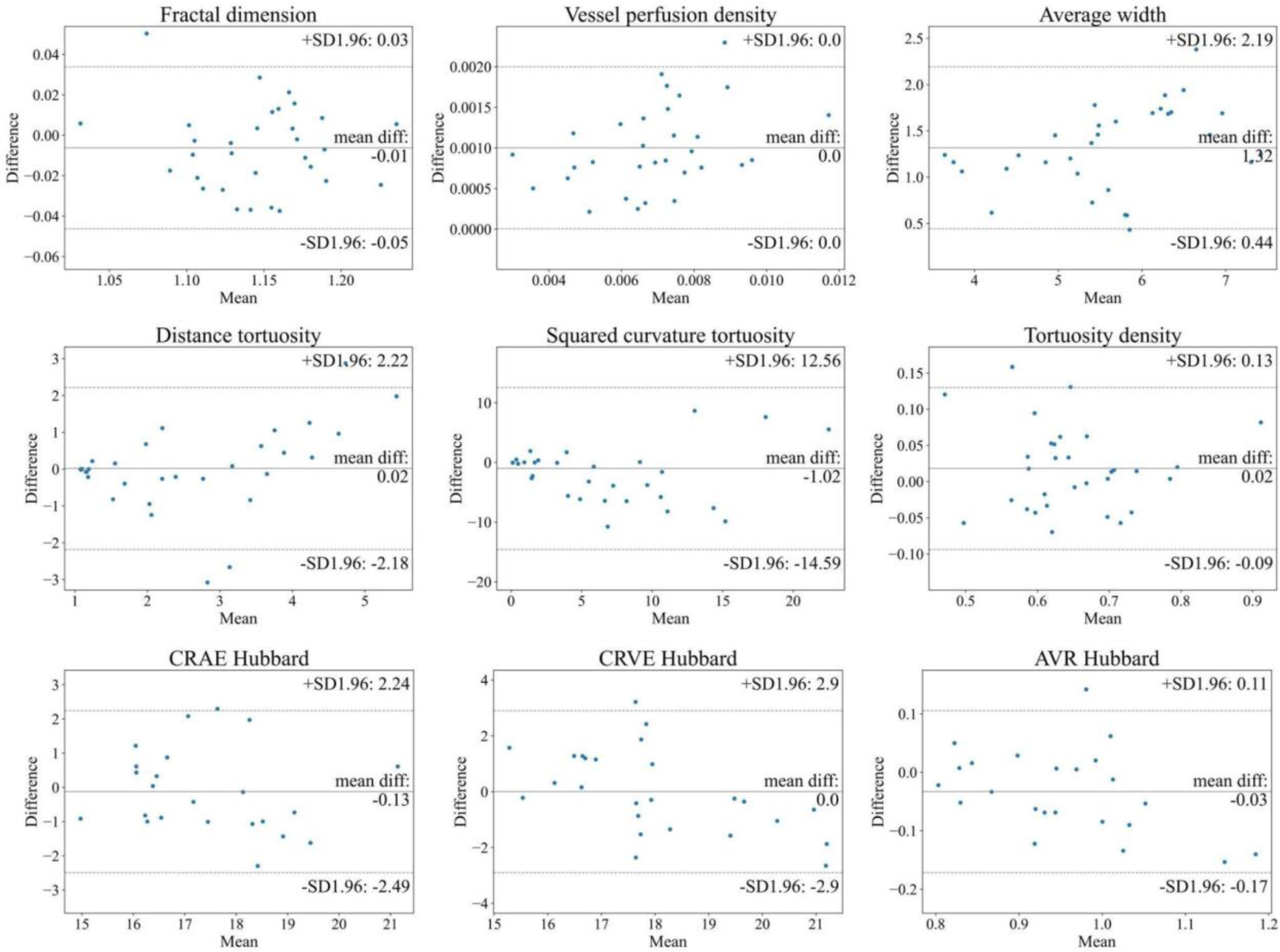
Bland-Altman plots of vascular feature agreement between expert annotation and AutoMorph segmentation at ZONE B. The first two rows features (tortuosity, fractal dimension, etc.) are calculated with binary vessel segmentation map from DR-HAGIS, while the last row features (calibre) are measured with artery/vein segmentation map from IOSTAR-AV. In each subplot, the central line indicates the mean difference and two dash lines represent 95% limits of agreement. The unit of average width, CRAE, and CRVE is pixel as resolution is unknown.

**Table 3.** Agreement calculation of measured vascular features between AutoMorph and expert annotation. The agreement of vessel calibre is validated on IOSTAR-AV dataset and other metrics are with DR-HAGIS dataset. Since calibre features rely on six largest arteries and veins in ZONE B and ZONE C, there is no calibre feature in the whole image level.

|  | ZONE B | ZONE C | Whole Image |
| --- | --- | --- | --- |
|  | ICC (95% CI) | ICC (95% CI) | ICC (95% CI) |
| DR-HAGIS |  |  |  |
| Fractal dimension | 0.94 (0.88-0.97) | 0.98 (0.95-0.99) | 0.94 (0.88-0.97) |
| Vessel density | 0.98 (0.96-0.99) | 0.97 (0.94-0.99) | 0.94 (0.88-0.97) |
| Average width | 0.95 (0.89-0.98) | 0.96 (0.93-0.98) | 0.97 (0.95-0.99) |
| Distance tortuosity | 0.80 (0.59-0.91) | 0.85 (0.69-0.93) | 0.86 (0.73-0.93) |
| Squared curvature tortuosity | 0.68 (0.34-0.85) | 0.88 (0.75-0.94) | 0.84 (0.68-0.92) |
| tortuosity density | 0.89 (0.77-0.95) | 0.70 (0.38-0.86) | 0.87 (0.74-0.93) |
| IOSTAR-AV |  |  |  |
| CRAE(Hubbard) | 0.81 (0.56-0.92) | 0.82 (0.57-0.91) | / |
| CRVE(Hubbard) | 0.8 (0.54-0.91) | 0.78 (0.52-0.89) | / |
| AVR(Hubbard) | 0.87 (0.69-0.94) | 0.81 (0.66-0.92) | / |
| CRAE(Knudtson ) | 0.76 (0.45-0.9) | 0.75 (0.44-0.89) | / |
| CRVE(Knudtson) | 0.85 (0.67-0.94) | 0.86 (0.58-0.9) | / |
| AVR(Knudtson) | 0.85 (0.66-0.94) | 0.82 (0.51-0.91) | / |

### Running efficiency and interface

The average running time for one image is about 20 seconds using a single GPU Tesla T4, from pre-processing to feature measurement. To ensure accessibility for researchers without coding experience, we make AutoMorph compatible with Google Colaboratory (free GPU) (Figure 7). The process involves placing images in a specified folder and then clicking the ‘run’ command. All results will be stored including segmentation maps and a file containing all measured features.

**Figure 7.**
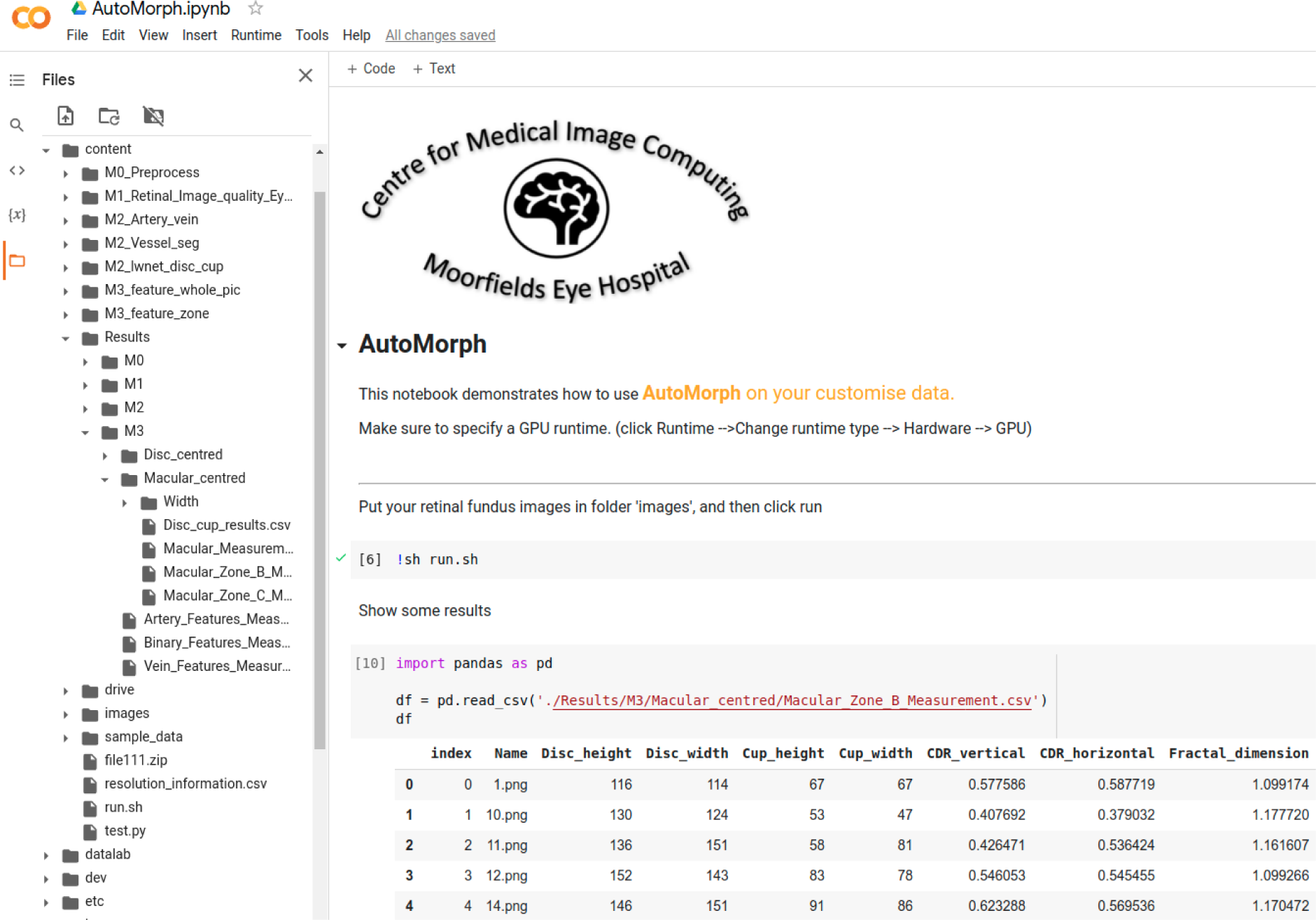
Interface of AutoMorph on Google Colab. After uploading images and clicking the ‘run’ button, all processes are executed and results stored, requiring no human intervention. Left side shows files directory and the right bottom lists five examples with parts of features.

## Discussion

In this report, the four functional modules of the AutoMorph pipeline achieve comparable, or better, performance compared with state-of-the-art for both image quality grading and anatomical segmentation. Furthermore, our approach to confidence analysis decreases the number of false gradable images by 76%, greatly enhancing the reliability of our pipeline. Hence, we learn that by a tailored combination of deep learning techniques, it is practical to accurately analyse the retinal vascular morphology in a fully automated way. Although we have evaluated the binary vessel segmentation model on ultra-widefield retinal fundus dataset AV-WIDE, we recommend using AutoMorph on retinal fundus photographs with a 25-60 field of view (FOV), as all the deep learning models are trained using images with FOV equals to 25-60 degree, and the pre-processing step is tailored for images with this FOV.

AutoMorph maintains computation transparency despite the use of deep learning techniques. Recently, similar systems have used deep learning models to skip intermediary steps and instead directly predict morphology features. For example, SIVA-DLS predicts vessel calibre from retinal fundus images without optic disc localisation or artery/vein segmentation ^3^. Another work directly predicts CVD factors from retinal fundus images in an end-to-end manner ^61^. Although these designs provide some insight into the applications of deep learning to ophthalmology, the end-to-end pipeline sacrifices transparency and raises interpretability concerns, representing a potential barrier to clinical implementation ^62, 63^. Specifically, considering that some formulas are empirically defined (e.g. CRAE and CRVE are calculated based on the six widest arteries and veins), it is hard to verify whether a model can learn this type of derivation. In contrast, the AutoMorph pipeline maintains transparency as the individual processes can be decomposed. Models are initially employed for anatomical segmentation, before vascular features are measured with traditional formulas. This process is consistent with the typical pipeline of human computation, thus improving the credibility of feature measurements.

The study cohort is selected by the image quality grading module. In this work, being different from previous work with only good quality images, we tried to explore the effectiveness of usable images. Although purely including good quality images can avoid potentially challenging cases for anatomic segmentation models (e.g., images with gloomy illumination), it filtered out the usable images which can contribute to a more general conclusion with a larger study cohort. Also, in clinical practice, a considerable number of images are usable quality but may not qualify as perfectly good quality. The pipeline developed in an environment similar to clinical reality is more prone to be deployed in the clinic. In image quality grading, the confidence analysis has recognised a considerable proportion of false gradable images and corrected them as reject quality by thresholding, as shown in Figure 3 and Figure 4. This avoids some reject quality images failing the anatomical segmentation and then generating large errors in feature measurement. Although this thresholding increased the false ungradable cases (green box in Figure 4(b)), the priority of recognising the false gradable images is secured. Of course, it is acceptable to only include the good quality images in the research cohorts, as the same as previous work, when the quantity of good quality images is large.

Although this work demonstrates the effectiveness of a deep learning pipeline for analysing retinal vascular morphology, there are some challenges remaining regarding technique and standardisation. Firstly, annotating retinal image quality is subjective and lacks strict guidelines, therefore it is hard to benchmark external validation performance. Secondly, there is still room for improving anatomical segmentation, especially for artery/vein segmentation. Thirdly, considering the agreement varies across various vascular features (Table 3), it is required to compare the robustness of these features and understand the pros and cons of each one. Finally, a uniform protocol for validating retinal analysis pipelines is required, considering existing software (RA ^28^, IVAN ^6^, SIVA ^29^, and VAMPIRE ^25^, etc.) shows high variation in feature measurement ^64, 65^. These four challenges exist in the field of ‘oculomics’, presenting an impediment to more extensive research.

We have made AutoMorph publicly available to benefit research in the field of ‘oculomics’ which studies the association between ocular biomarkers and systemic disease. We have designed

AutoMorph’s interface using Google Colaboratory to facilitate its use by clinicians without coding experience. In future work, we will investigate the solution dedicated to the above challenges in ‘oculomics’ research. Also, the feasibility of automatic pipeline can be extended to other modalities, e.g., optical coherence tomography (OCT) and OCT angiography.

## Data Availability

All data produced are available online

## Supplementary material: Feasibility of Analysing Retinal Vascular Morphology via a Deep Learning Pipeline

### S1. Datasets

#### Dataset and ground truth

All the datasets for model training and external validation are publicly available. Each dataset consists of a number of retinal fundus photographs and corresponding ground truth. We summarise the characteristic of each dataset as well as the information of the ground truth.

Image quality grading datasets include EyePACS-Q and DDR-test. EyePACS-Q is based on EyePACS, which contains a large set of images collected by different imaging devices during diabetic retinopathy screening. During EyePACS-Q construction, two experts were asked to grade the image quality into three categories: good, usable, and reject. Good quality images have no low-quality factors, such as uneven illumination and blur, and all retinopathy characteristics are clearly visible. Usable quality images show some slight low-quality factors, but the optic disc, macula region, and lesions are clear enough to be identified by ophthalmologists. Reject quality has a serious quality issue and cannot be used to provide a full and reliable diagnosis, even by ophthalmologists 31. For the DDR-test dataset, there are two categories for image quality grading, gradable and ungradable. Images with a blurring area of more than 70% and without clearly visible diabetic retinopathy lesions are considered ungradable. Seven professional graders have been trained by ophthalmologists. The seven graders vote to evaluate the images in question, and the final classification was determined by majority voting. If the seven graders could not determine a classification result, they will consult more experienced experts 32. The EyePACS-Q and DDR datasets indeed establish the ground truth based on the expert assessment of the image’s gradability/diagnosability. This can help identify retinal fundus photographs with serious artefacts, uneven illumination, and low contrast, which are also challenging cases for the deep learning segmentation methods (anatomical segmentation modules). In this case, by filtering the ungradable images, we can get a more accurate segmentation map as well as a precise vascular feature estimation.

For binary vessel segmentation, the ground truth map has the same size as the retinal fundus photographs. The vessel pixels are white colour and the background pixels are black. The DRIVE 33 dataset consists of a total of 40 retinal fundus photographs, including 7 diabetic retinopathy lesions. These images were labelled by an ophthalmological expert. For the STARE 34 dataset, ten of the images are of patients with no pathology and ten of the images contain pathology that obscures or confuses the blood vessel appearance in varying portions of the image. CHASEDB1 ^35^ is acquired from multiethnic school children, being a part of a cardiovascular health survey in 200 primary schools in London, Birmingham, and Leicester, and labelled by two experts. The ground truth of the HRF ^36^ dataset is generated by a group of experts working in the field of retinal image analysis and clinicians from the cooperated ophthalmology clinics. The IOSTAR ^37^ dataset is annotated by a group of experts working in the field of retinal image analysis. The LES-AV ^38^ comprises 22 fundus photographs with available manual segmentations of the retinal vessels and their expert classification into arteries and veins. The external validation data includes DR-HAGIS ^40^ and AV-WIDE ^19, 39^. DR-HAGIS has 39 fundus images with four subgroups, glaucoma, hypertension, diabetic retinopathy, and age-related macular degeneration. The manual segmentation is provided by an expert grader. AV-WIDE provides the graph-based annotations by an expert ophthalmologist and AV-WIDE is a widefield retinal fundus dataset.

The artery/vein ground truth map is characterised by four colours: red for arteries, blue for veins, green for uncertain pixels (unidentifiable vessels at intersections of arteries and veins), and black for the background. We use three training datasets, the DRIVE-AV 33,41, LES-AV 38, and HRF-AV 36,42, and one external validation dataset IOSTAR-AV. The DRIVE-AV dataset supplements the artery/vein ground truth by an expert to the DRIVE dataset. HRF-AV’s initial labelling was carried out by an expert in retinal image analysis, and then carefully corrected by an ophthalmologist. IOSTAR-AV 37,43 is annotated by a group of experts working in the field of retinal image analysis.

Optic disc segmentation is a binary segmentation task that classifies each pixel as the optic disc pixel or the background. The ground truth includes the white colour pixels for the optic disc and black colour for the background. For the REFUGE dataset, the ground truths of the optic disc were provided by seven independent glaucoma specialists from the Zhongshan Ophthalmic Center (Sun Yat-sen University, China). All the ophthalmologists independently reviewed and a single segmentation per image was afterwards obtained by taking the majority voting of the annotations of the seven experts.

A senior specialist with more than 10 years of experience in glaucoma performed a quality check afterwards, analyzing the resulting masks to account for potential mistakes 44. For the GAMMA dataset, four clinical ophthalmologists manually annotated the initial segmentation region of the optic disc for each fundus image. The senior ophthalmologist then fused the results of the four initial segmentation results and selected the intersection of the annotated results of several ophthalmologists as the final ground truth 46,45. For the external validation dataset IDRID, all observers were trained by expert ophthalmologists for the identification of individual lesions and optic discs. Later the markings on each of these images were reviewed by two retinal specialists, and they were finalized when the necessary consensus was reached 47.

Some examples are shown in Figure S1 to depict the visualisation difference brought by different imaging devices and research studies.

**Figure S1.**
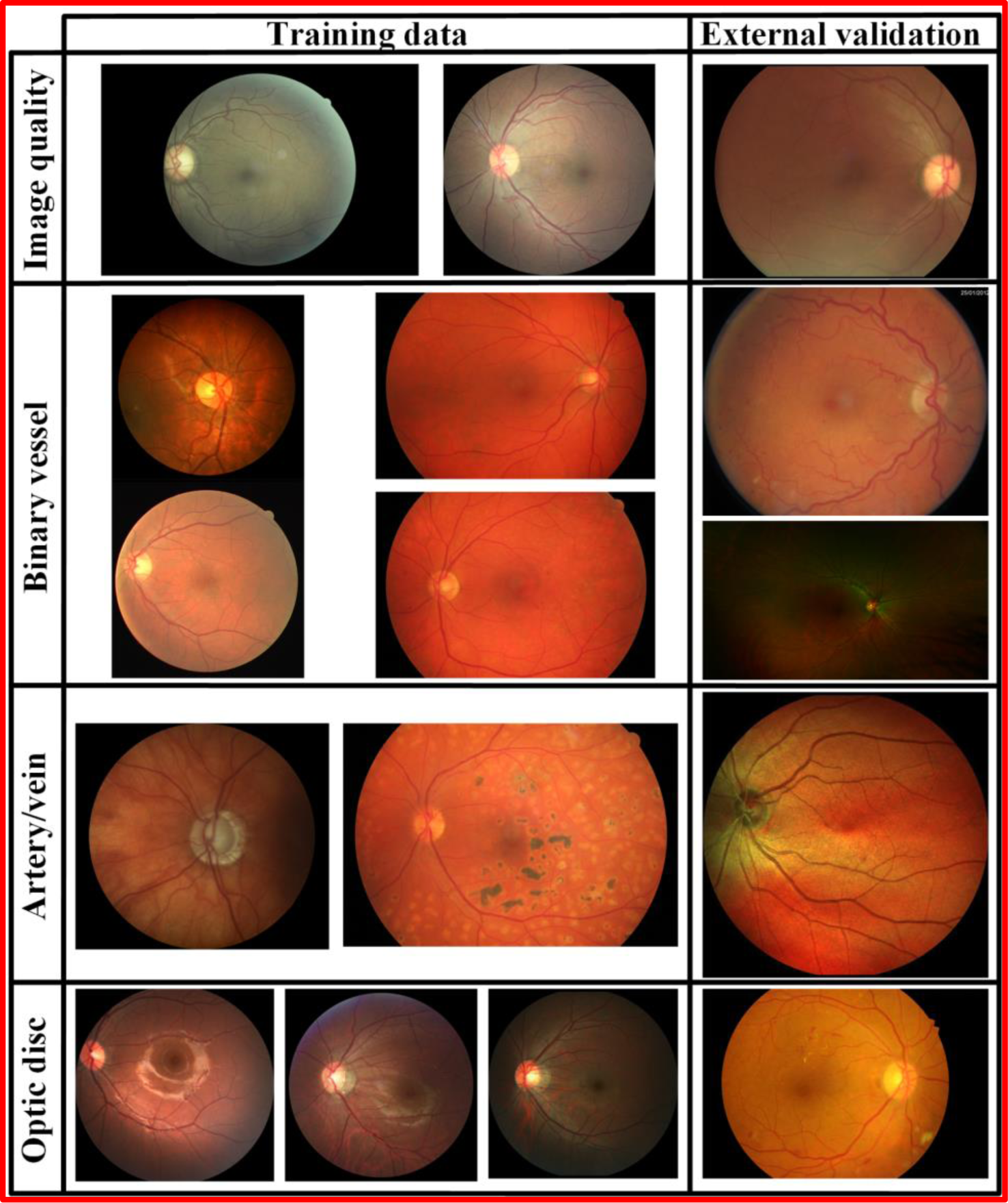
Image examples of training data and external validation data for the image quality grading, binary vessel segmentation, artery/vein segmentation, and optic disc segmentation.

### S2. Image quality grading module Image pre-processing

The images are squared as Figure S2.

**Figure S2.**
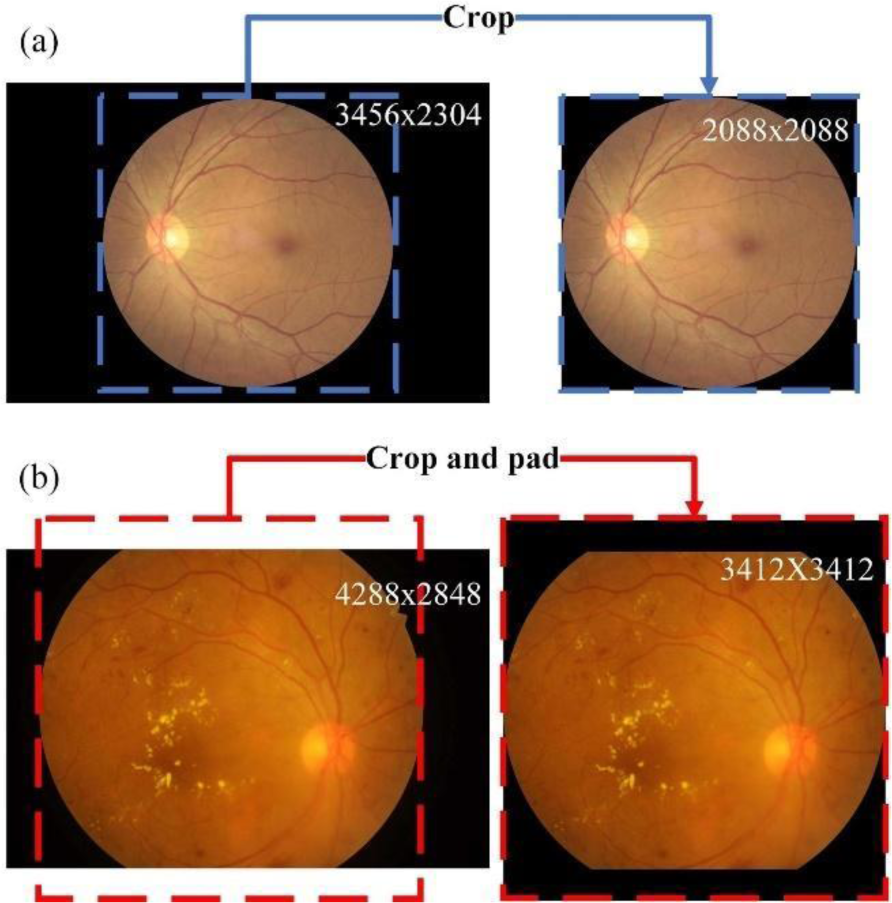
Two examples of retinal fundus image pre-processing. The redundant background is removed by cropping in (a) and (b), whilst some padding is employed in (b) to make the image conform to a geometric shape. The image size is listed at the top right corner.

#### Model structure selection

We compared the classification performance with different classification backbones, namely ResNeXt101-32x8d, EfficientNet-B4, and EfficientNet-B5. The models are pretrained on the ImageNet dataset, and the final classifier neurons are substituted from 1000 to 3, respectively corresponding to the category of good, usable, and reject image quality. The training epoch is 30 and batch size is 8. The initial learning rate is 0.0002 and the optimiser is Adam. The learning rate schedule and early stopping are employed to avoid model overfitting. The cross entropy loss function is used and the checkpoint with lowest loss is stored for inference. We use four Tesla T4 GPUs to train the model. As shown in Figure S3, the performance of the three backbones shows no statistical difference (p>0.05). In this case, we use the EfficientNet-b4 as the backbone considering its lower parameters compared with EfficientNet-b5 and ResNeXt101-32x8d.

**Figure S3.**
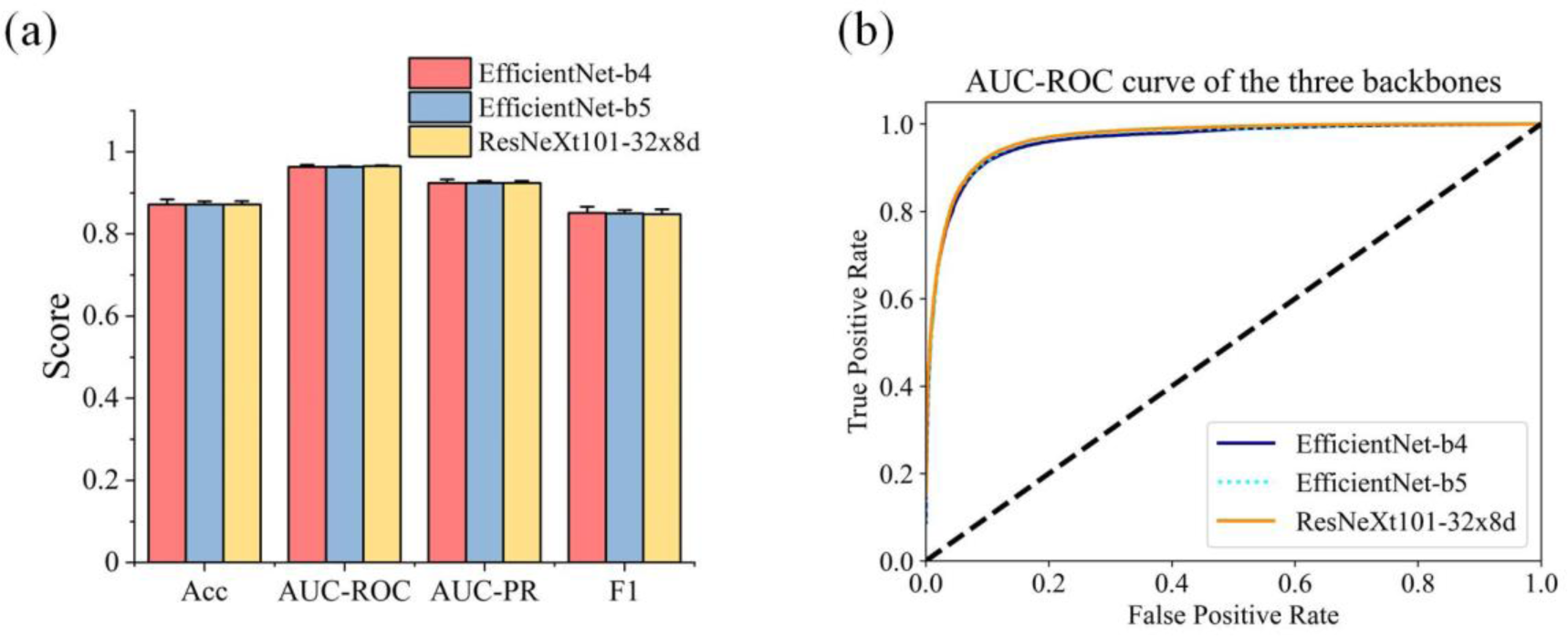
Performance comparison of the quality grading models with different backbones. (a) verifies there is no significant difference between the three backbones and (b) shows that the AUC-ROC curves highly overlap. For each backbone, eight models are trained on different subsets of EyePACS-Q training data, and then validated on EyePACS-Q validation data. Acc, accuracy. AUC-ROC, Area-under-curve Receiver operating characteristic. AUC-PR, Area-under-curve Precision-Recall. F1, F1-score.

#### Confidence analysis threshold

We calculate the mean value and SD of ensemble probability in confidence analysis. Depicted in Figure S4, comparing the distribution of true gradable and false gradable in the tuning data (20% of training data used to tune the training hyperparameters during training), we found the FP cases have lower mean value and higher SD. Threshold choices were made by inspecting average probability histograms and standard deviation histograms. We selected thresholds that resulted in a desirable trade-off, i.e., reduced a large proportion of false gradables whilst introducing an acceptable number of false ungradables. The average threshold of 0.75 and SD threshold of 0.1 are employed.

**Figure S4.**
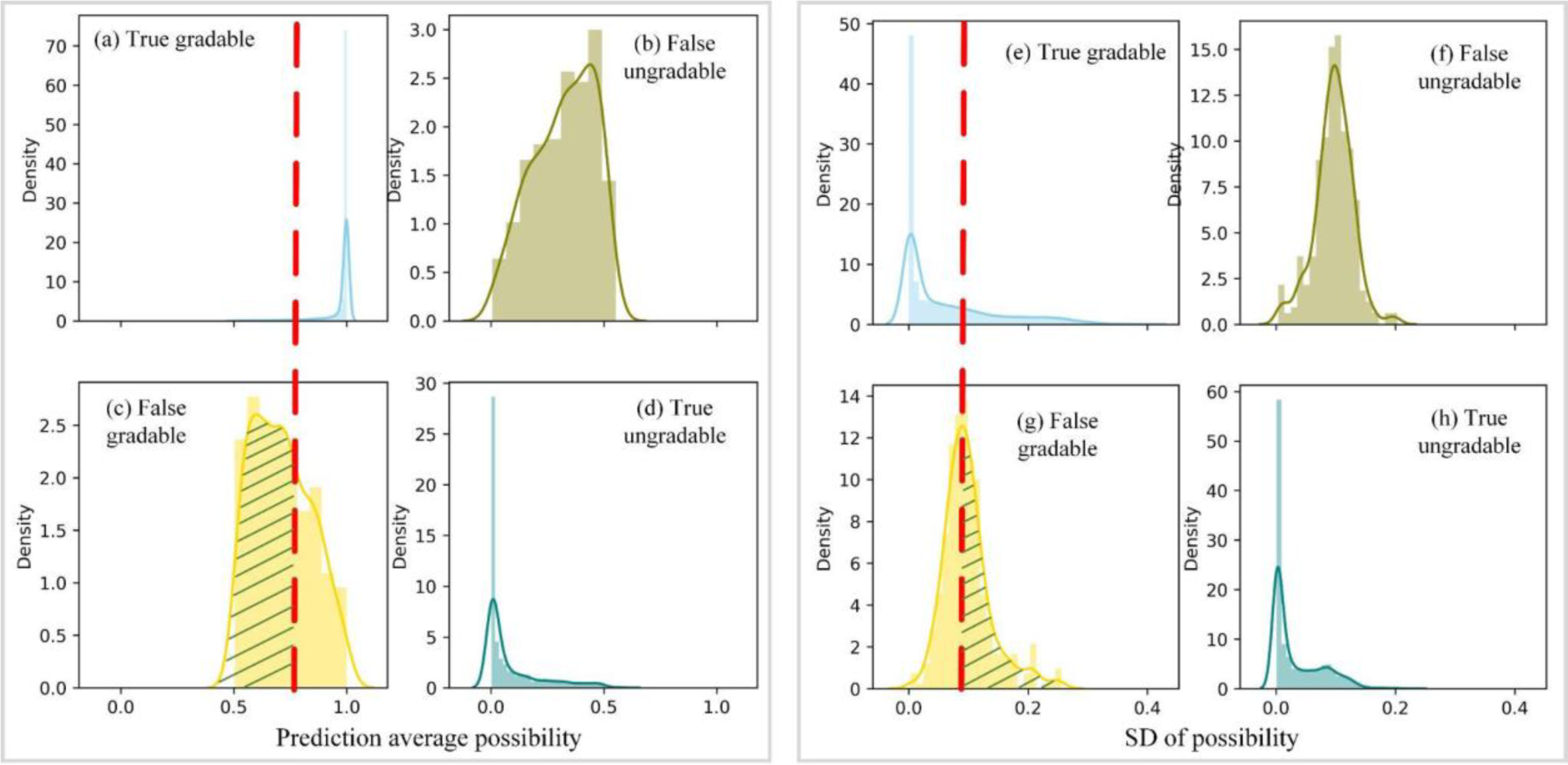
(a-d) show the average probability histogram, respectively indicating true gradable, false ungradable, false gradable, and true ungradable. (e-h) are corresponding SD histograms. Comparing the true gradable and false gradable, it’s observed that the average probability of true gradable (a) concentrates to 1 and SD (e) is closer to 0. By thresholding on an average probability 0.75 and SD threshold 0.1 (red dash line), lots of false gradable images can be rectified as the ungradable images and then being filtered out, depicted in regions with green slash lines.

#### Confusion matrix on DDR

The confusion matrix and AUC-ROC curve on DDR are listed in Figure S5.

**Figure S5.**
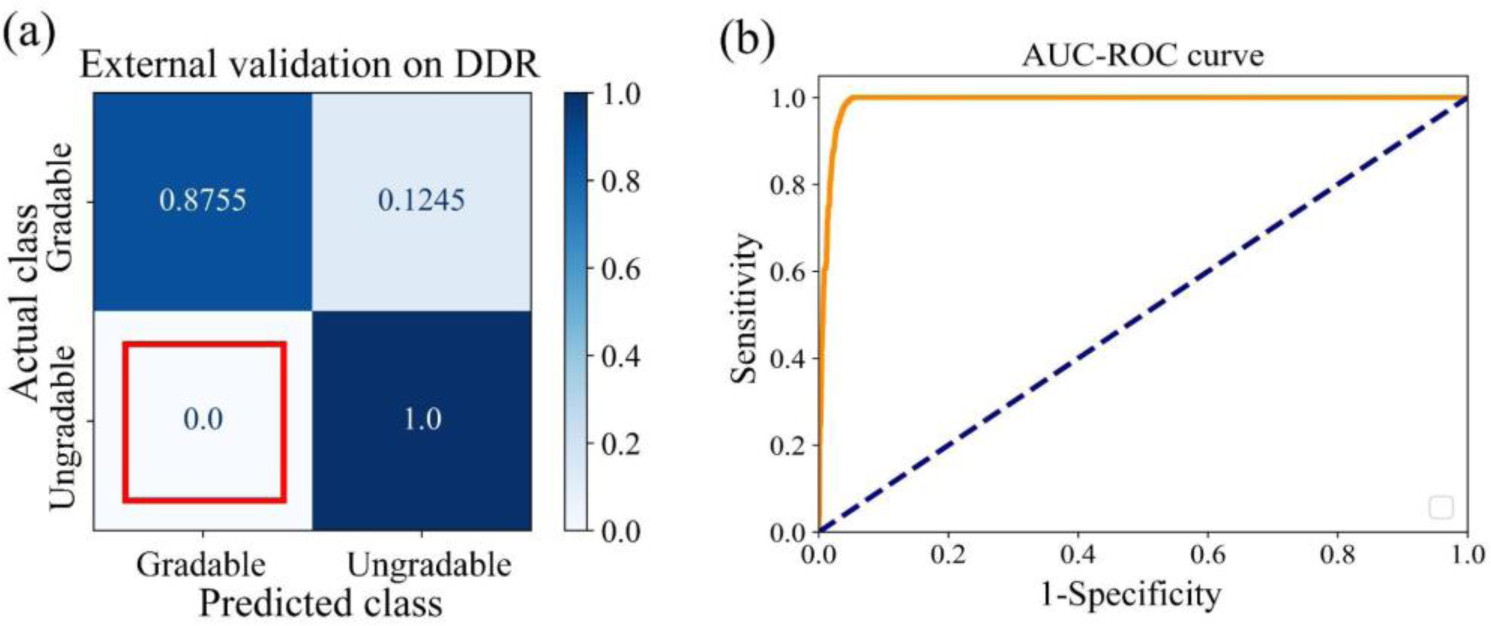
The confusion matrix (a) and AUC-ROC curve (b) showing the performance of AutoMorph on the external DDR test. The ratio of FP-gradable is 0 which says all ungradable images are correctly recognised, thus remaining no false gradable images for the following anatomical segmentation steps. The AUC-ROC curve indicates good performance of AutoMorph on the DDR test data.

### S3. Anatomical segmentation modules

#### Binary vessel segmentation

SEGAN is a variant of U-Net with enhanced performance in multi-scale information learning. It consists of a segmenter and a discriminator, which are trained in adversarial learning strategy. The input of the network is the retinal fundus photographs and the output is a segmentation map with the same size of the input. The pre-processed images are resized to (912, 912) to ease the computational stress. The segmentation map will be resized to the original size for feature measurement. Each pixel of the segmentation map shows the probability of being vessel. After thresholding with 0.5, the pixels with values of 1 are vessels. The training epoch is 600 and batch size is 2. The initial learning rate is 0.0002 and the optimiser is Adam. The learning rate schedule is employed for robust convergence. Three loss functions are utilised, namely adversarial loss, mean square error, and cross entropy loss. The checkpoint with the highest F1-score is stored. We use one Tesla T4 GPUs to train the model.

#### Artery/vein segmentation

BFN decomposes the multi-class task into multi binary tasks, followed by a binary-to-multi-class information fusion, so as to correct the information around the intersections. Different segmenters share the same discriminator. The input of the network is the retinal fundus photographs and the output is a multi-class segmentation map with the same size of the input. The pre-processed images are resized to (720, 720) to ease the computational stress. The multi-class segmentation map will be resized to the original size for feature measurement. Each pixel of the segmentation map shows the four-element probability of being artery, vein, uncertain pixels, and background. After selecting the maximised probability as the final category, the pixels with values of 0 are background, 1 for arteries, 2 for veins, and 3 for uncertain pixels. The red, blue, green, and black colours are transformed visualisation. The training epoch is 1500 and batch size is 2. The initial learning rate is 0.0008 and the optimiser is Adam. The learning rate schedule is employed. Three loss functions are utilised, namely adversarial loss, mean square error, and cross entropy loss. The checkpoint with the highest F1-score is stored. One Tesla T4 GPU is employed to train the model.

**Figure S6.**
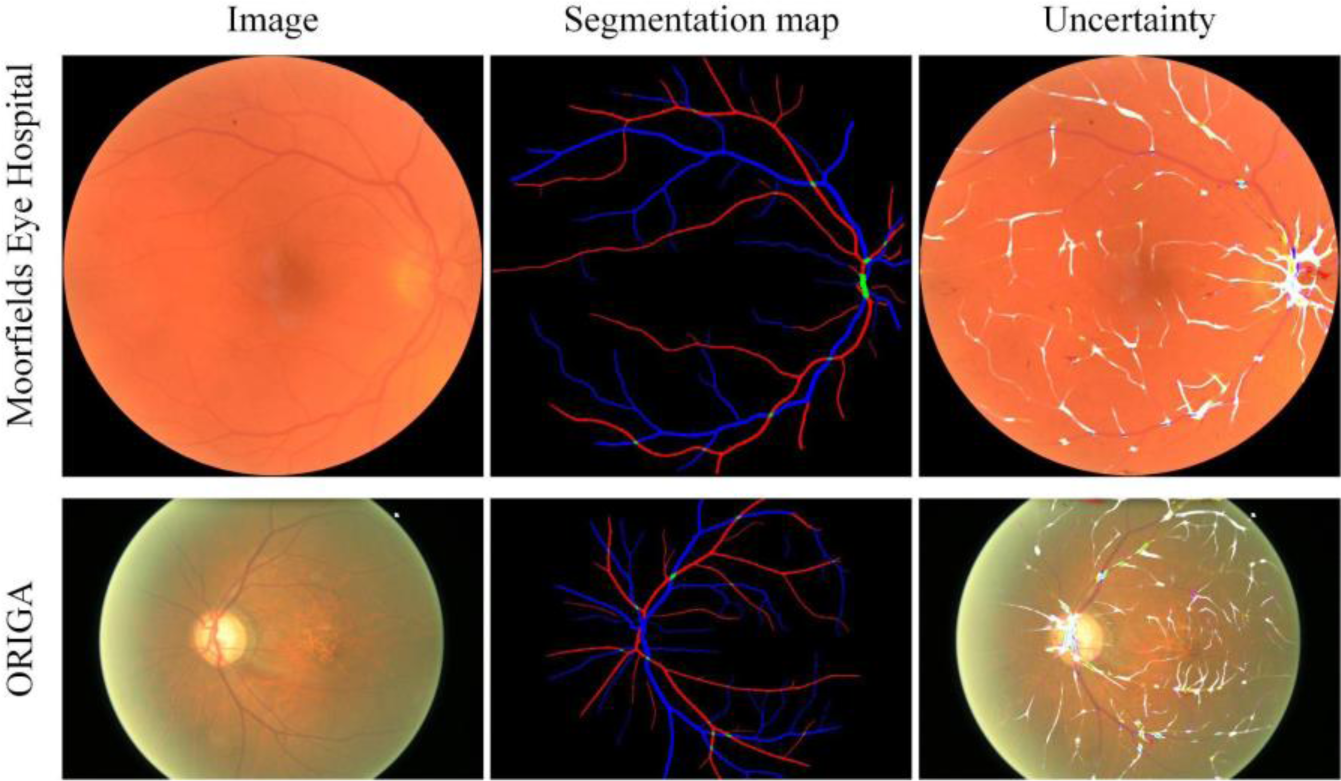
Artery/vein segmentation visualisation examples of challenging cases (blur and poor illumination) from Moorfields Eye Hospital and ORIGA dataset. The uncertainty map is overlaid on the retinal fundus image. Brighter white indicates higher uncertainty.

#### Optic disc segmentation

LW-Net consists of two simplified U-Net, where the first part generates a coarse segmentation result and the second part polishes the segmentation detail. This cooperated regularisation between the two parts enhances model segmentation performance when pathology is present. The overall model is light-weight in terms of parameter numbers. The input of the network is the retinal fundus photographs and the output is a segmentation map with the same size of the input. The pre-processed images are resized to (512, 512) to enable a large batch size. The segmentation map will be resized to the original size for feature measurement and the definition of the pre-defined zones. Each pixel of the segmentation map shows the probability of being optic disc. After thresholding with 0.5, the pixels with values of 1 are optic disc. The training epoch is 1000 and batch size is 16. The initial learning rate is 0.01 and the optimiser is Adam. The learning rate schedule is employed. The cross entropy loss is used and the checkpoint with the highest AUC-ROC is stored.

One Tesla T4 GPU is employed to train the model

In order to give a direct technical comparison between our segmentation methods and those compared methods, we have listed the technically fair comparison in Table S1. All models are trained on the same training dataset and internally validated. As we don’t have multiple groups of results for the compared methods ^39, 57–59^, we cannot test if a statistically significant difference exists. But our results show clearly better performance. We compare all the metrics they have provided. We follow the same splitting of training and testing data of the compared methods.

**Table S1.** Internal validation of AutoMorph and comparison to competitive methods. All models are trained and validated on the same data to compare the technical superiority.

Internal validation on **DR-HAGIS** (binary vessel segmentation)
|  | Sensitivity | Specificity | F1-score | Accuracy |
| --- | --- | --- | --- | --- |
| AutoMorph | 0.74±0.02 | 0.99±0.01 | 0.77±0.01 | 0.97±0.01 |
| Compared <sup>57</sup> | 0.67 | 0.98 | 0.71 | 0.97 |

Internal validation on **WIDE** (binary vessel segmentation)
|  | Sensitivity | Precision | F1-score | Accuracy |
| --- | --- | --- | --- | --- |
| AutoMorph | 0.81±0.02 | 0.85±0.02 | 0.83±0.02 | 0.98±0.01 |
| Compared <sup>39,57</sup> | 0.78 | 0.82 | 0.8 | 0.97 |

Internal validation on **IOSTAR-AV** (artery/vein segmentation)
|  | Sensitivity | Specificity | Accuracy |
| --- | --- | --- | --- |
| AutoMorph | 0.74±0.04 | 0.98±0.01 | 0.97±0.02 |
| Compared <sup>58</sup> | 0.79 | 0.76 | 0.78 |

Internal validation on **IDRID** (optic disc segmentation)
|  | Sensitivity | Accuracy | IoU |
| --- | --- | --- | --- |
| AutoMorph | 0.95±0.03 | 0.99±0.01 | 0.93±0.02 |
| Compared <sup>58,59</sup> | 0.9 | 0.99 | 0.85 |

### S4. Vascular feature in ZONE C and whole image

Bland-Altman plots for parts of vascular morphology features at ZONE B and whole image.

**Figure S7.**
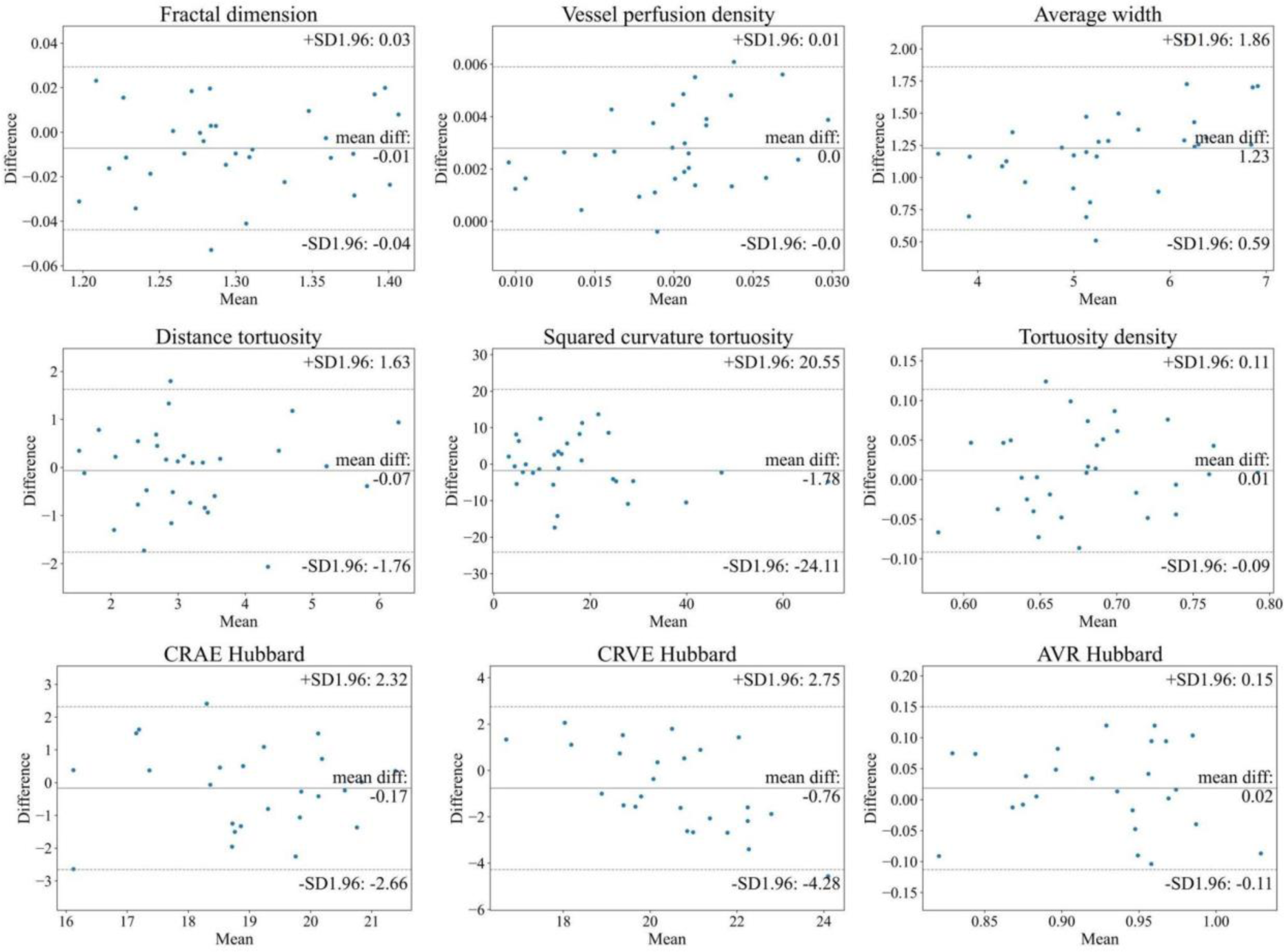
Bland-Altman plots of vascular feature agreement between expert annotation segmentation map and AutoMorph segmentation at ZONE C. The first two rows features (tortuosity, fractal dimension, etc.) are calculated with binary vessel segmentation map from DR-HAGIS, while the last row features (calibre) are measured with artery/vein segmentation map from IOSTAR-AV. In each subplot, the central line indicates the mean difference and two dash lines represent 95% limits of agreement.

**Figure S8.**
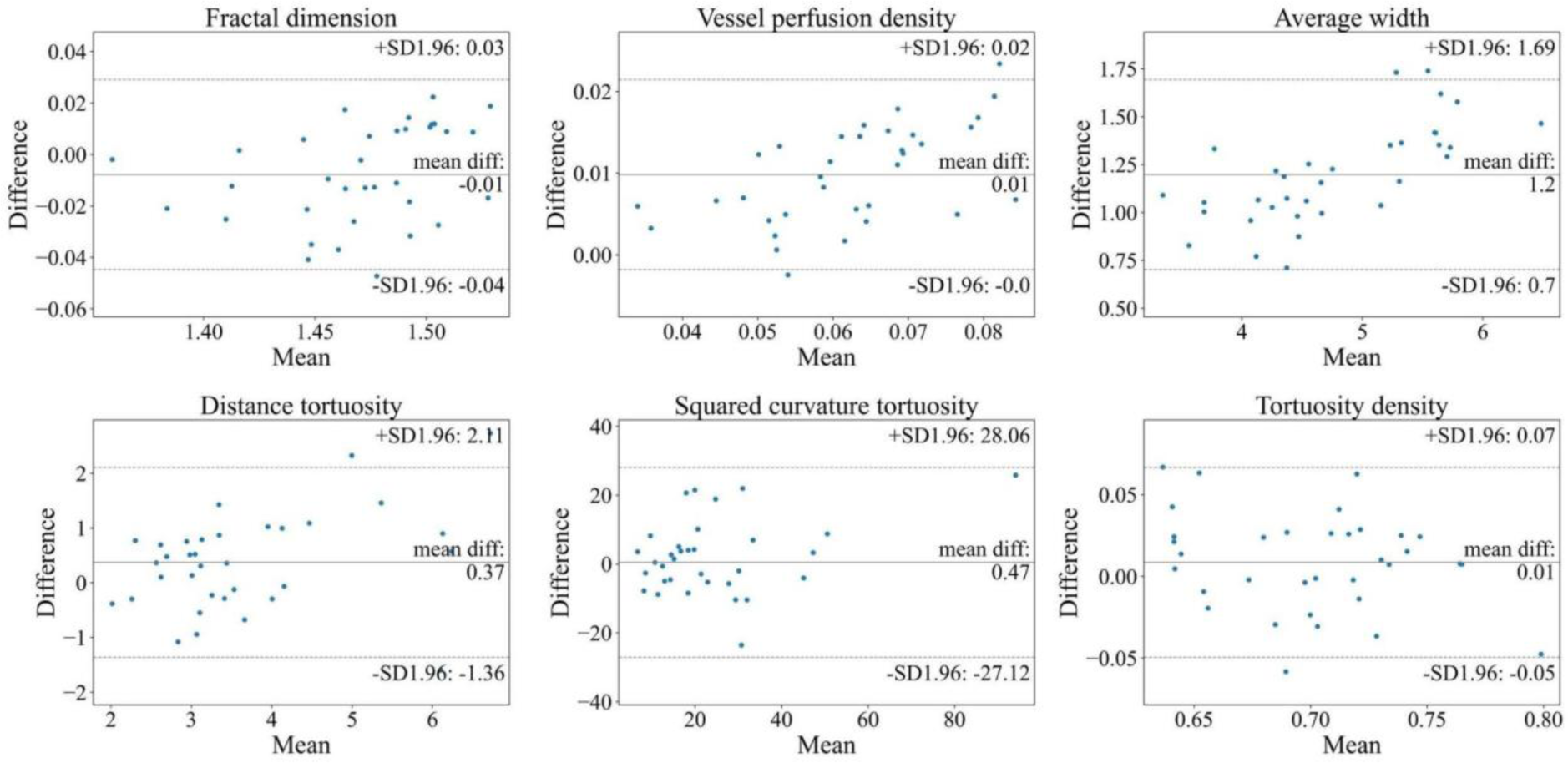
Bland-Altman plots of vascular feature agreement between expert annotation segmentation map and AutoMorph segmentation at the whole image.

**Figure S9.**
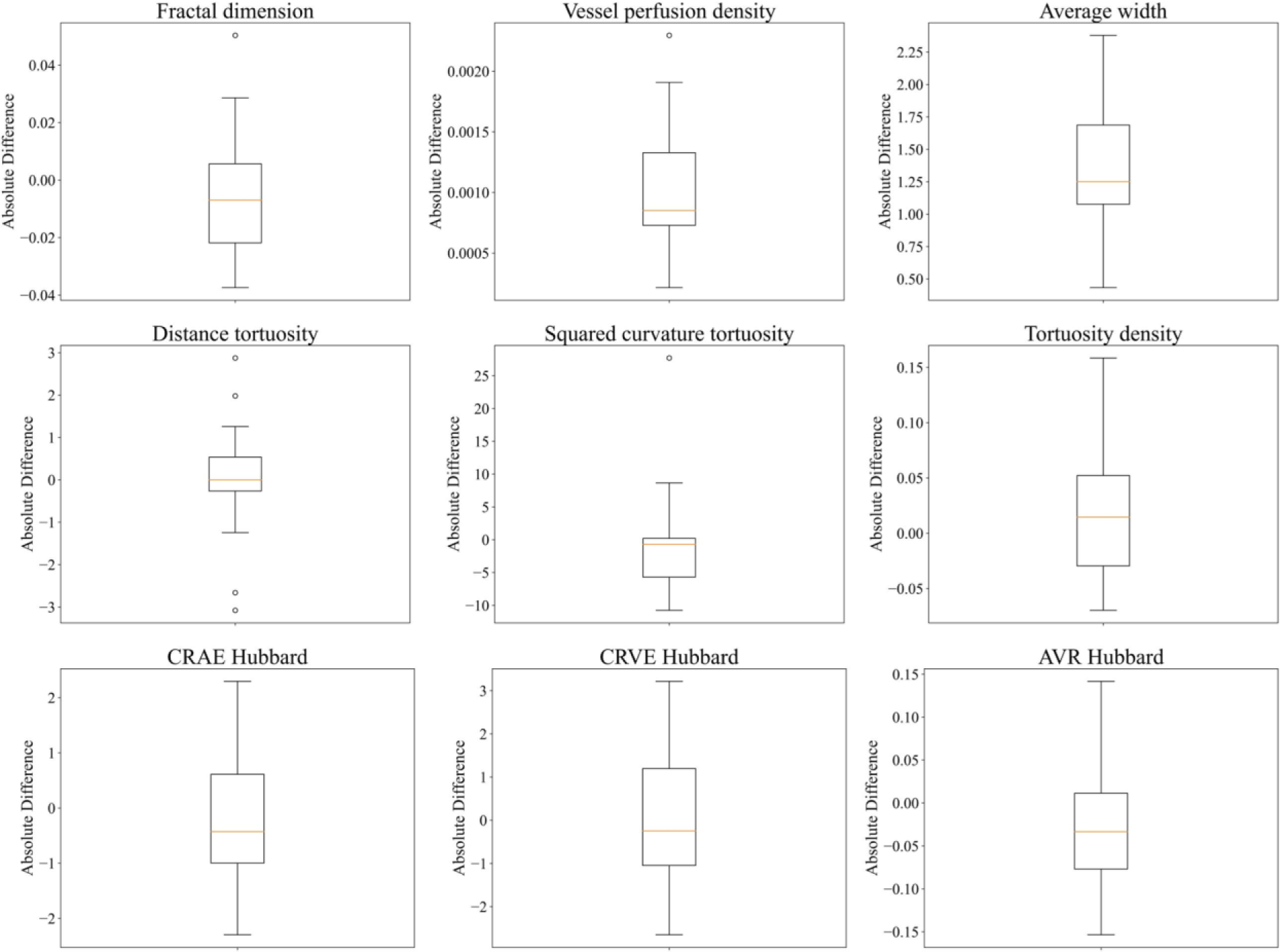
Boxplot of vascular feature difference between expert annotation segmentation map and AutoMorph segmentation at the ZONE B. The first two rows features (tortuosity, fractal dimension, etc.) are calculated with binary vessel segmentation map from DR-HAGIS, while the last row features (calibre) are measured with artery/vein segmentation map from IOSTAR-AV. In each subplot, the orange line indicates the mean difference and the box includes data distribution between 25 percentile to 75 percentile. Circles are outliers.

**Figure S10.**
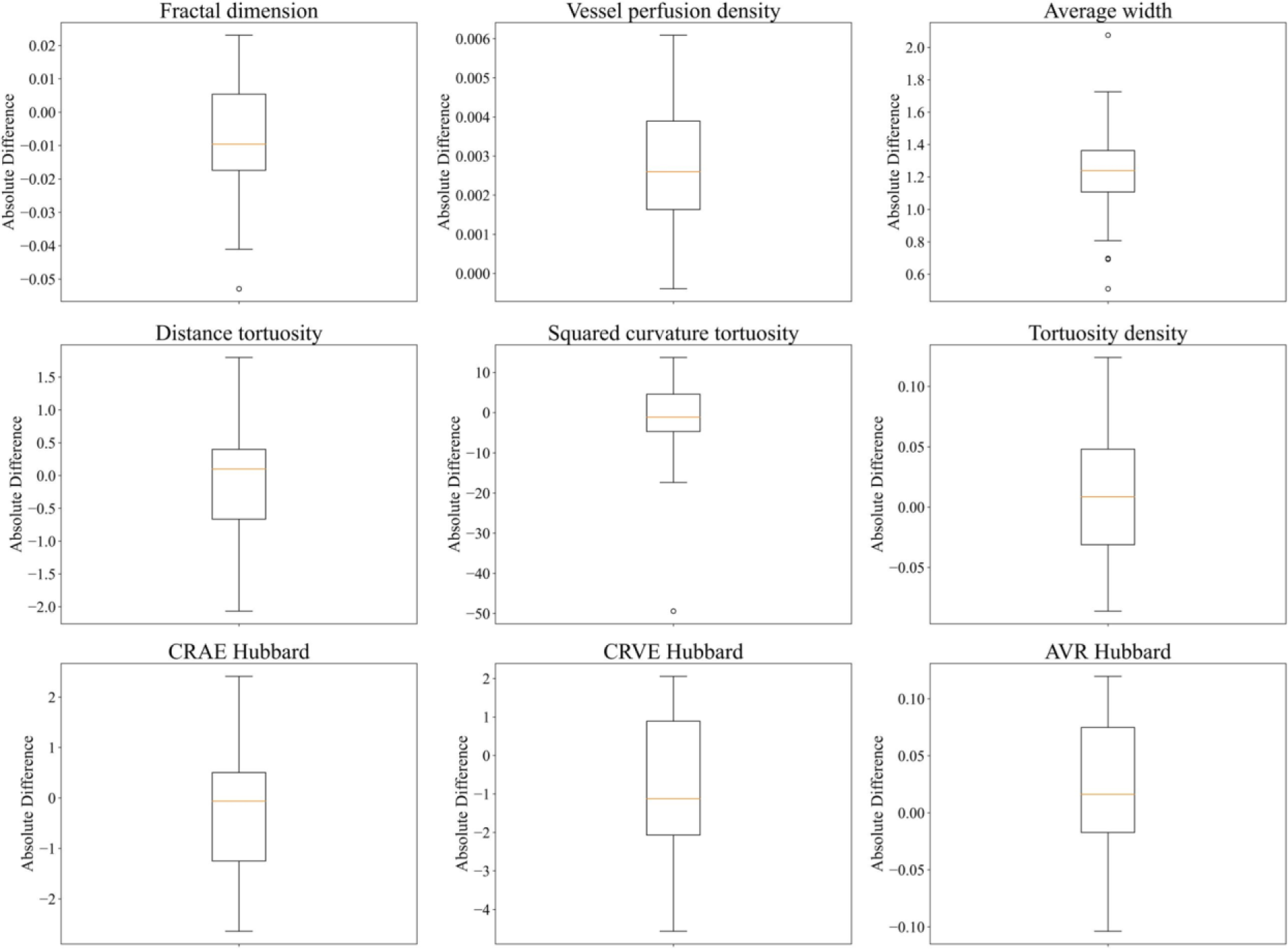
Boxplot of vascular feature difference between expert annotation segmentation map and AutoMorph segmentation at the ZONE C.

**Figure S11.**
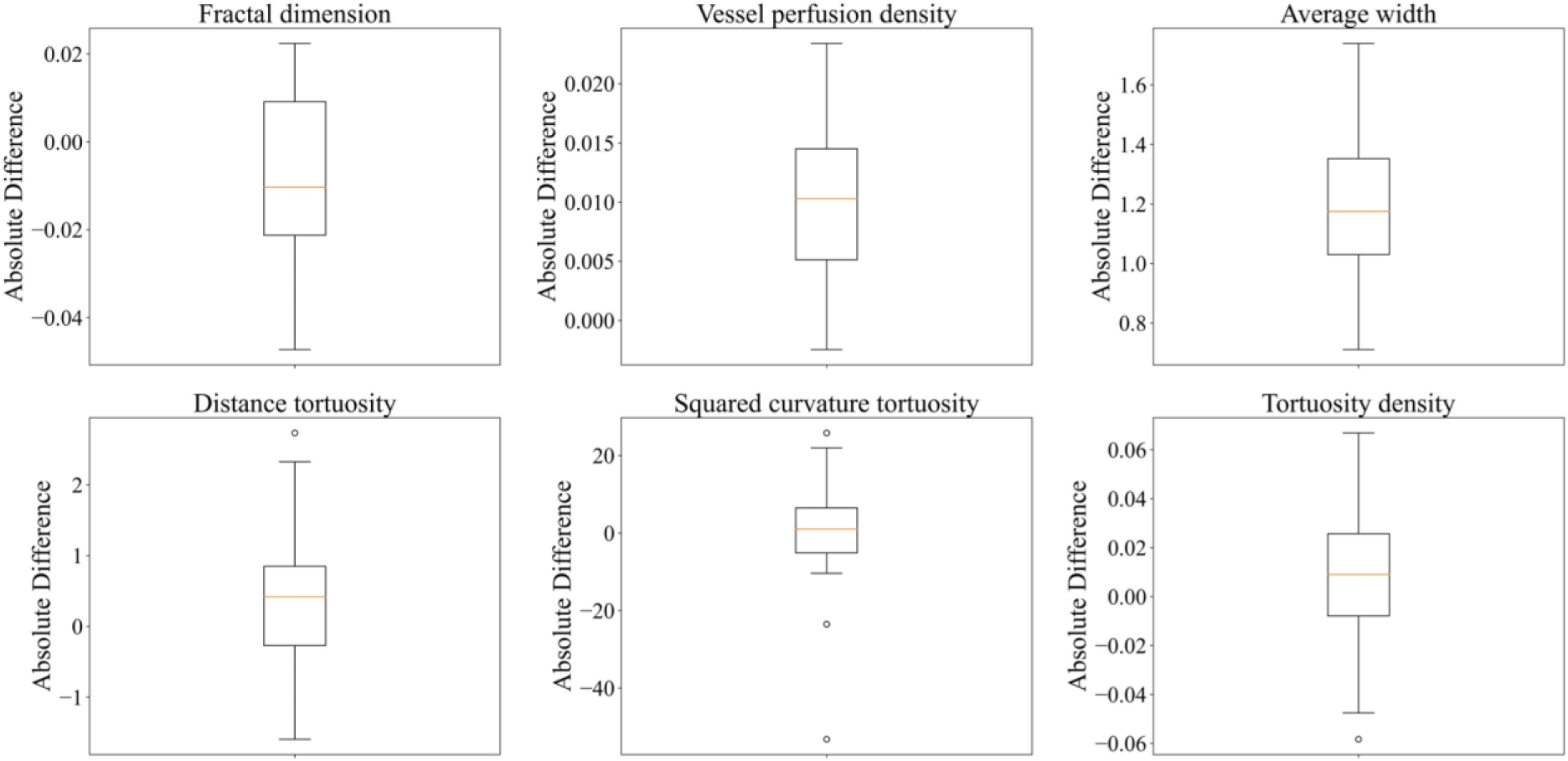
Boxplot of vascular feature difference between expert annotation segmentation map and AutoMorph segmentation at the whole image.

**Figure S12.**
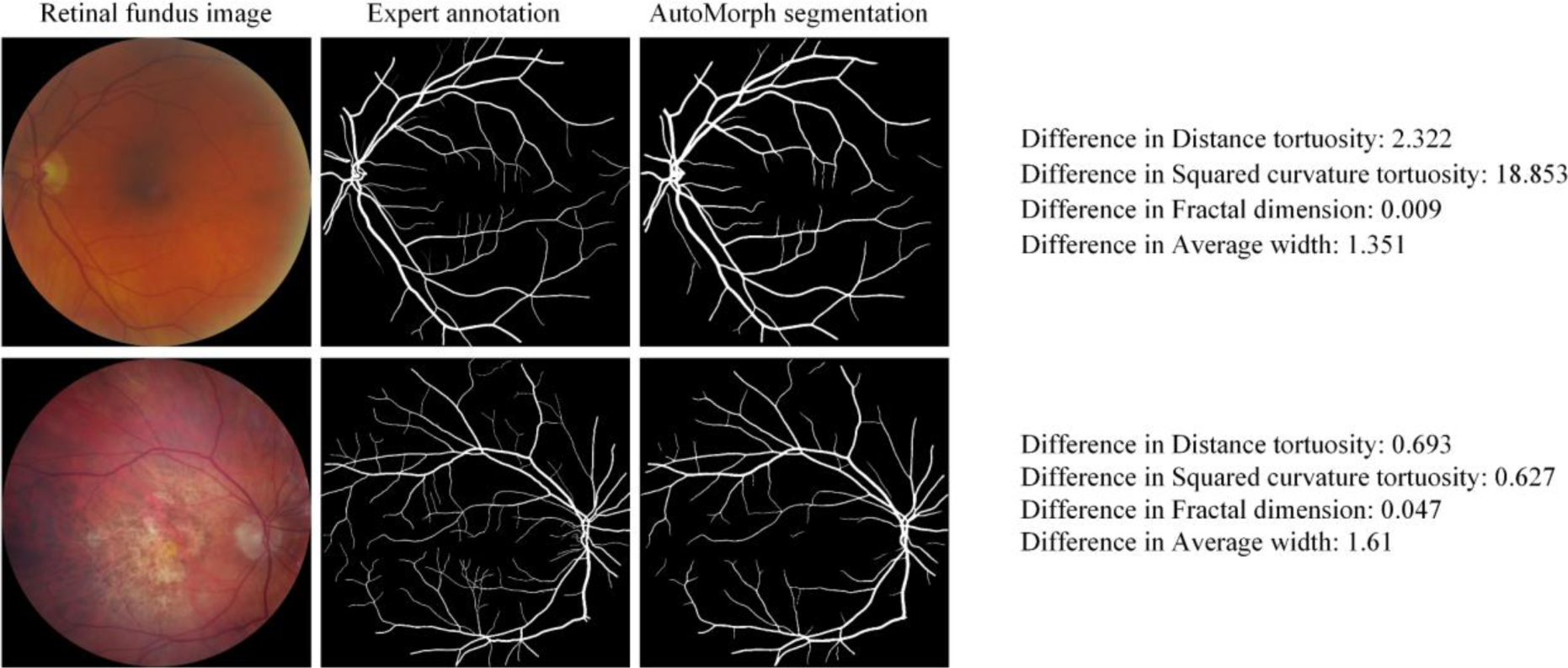
Examples with large vascular features’ error. Comparing the two cases, we can understand that different vascular features are sensitive to specific kinds of segmentation error according to the calculation process. Distance tortuosity and squared curvature tortuosity usually show large errors when vessels show extra junctions or miss junctions, while the fractal dimension shows errors when overall vasculature shape is considerably different to expert annotation (e.g., missing some distal vessels).

### S5. Detailed list of measured features

All measured features are listed in Figure S13.

**Figure S13.**
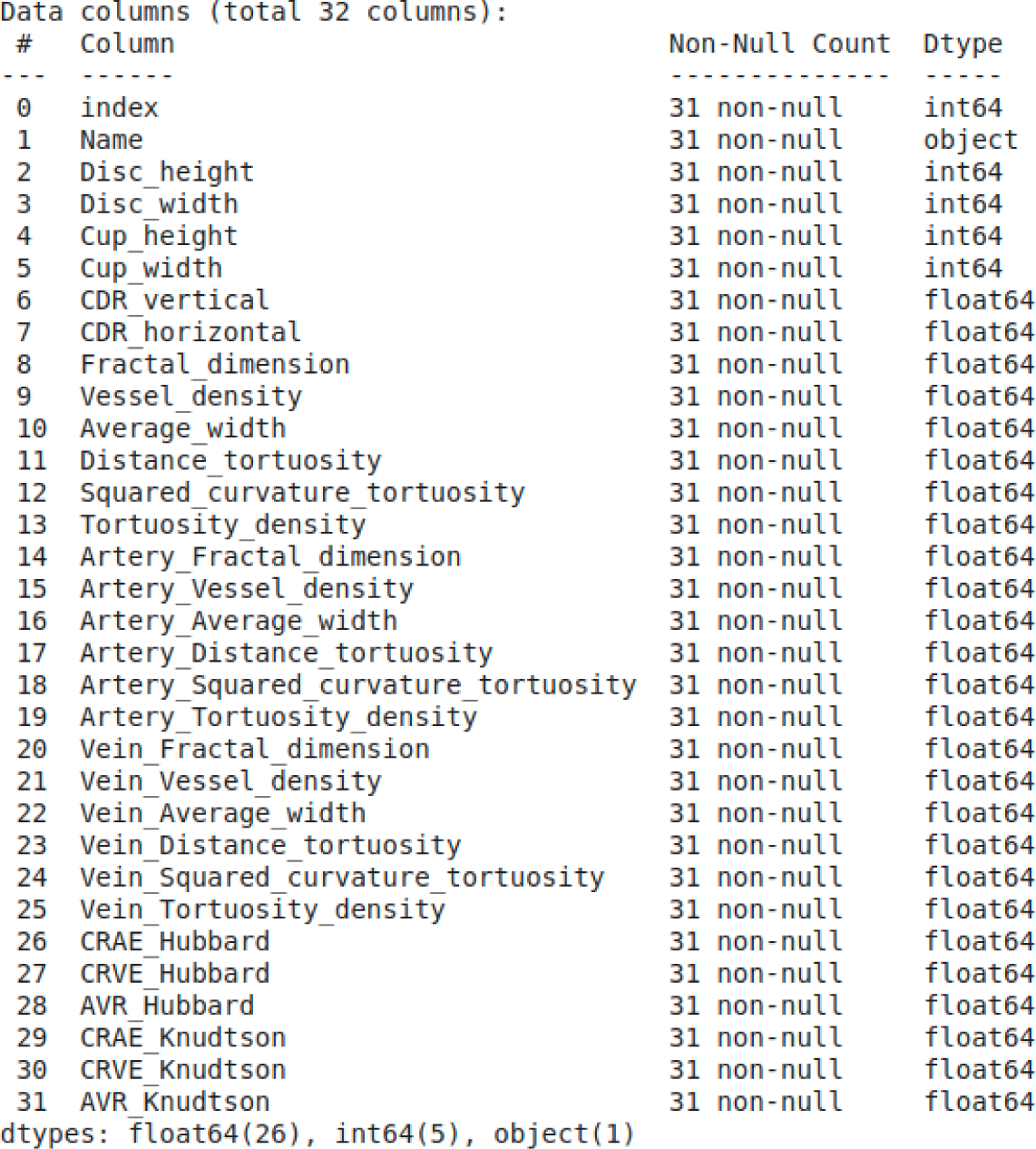
This is a comprehensive list for measured vascular features.

